# Afucosylation of HLA-specific IgG1 as a potential predictor of antibody pathogenicity in kidney transplantation

**DOI:** 10.1101/2022.03.09.22272152

**Authors:** Pranay Bharadwaj, Sweta Shrestha, Tamas Pongracz, Catalano Concetta, Shilpee Sharma, Alain Le Moine, Noortje de Haan, Naoka Murakami, Leonardo V. Riella, Vanda Holovska, Manfred Wuhrer, Arnaud Marchant, Margaret E. Ackerman

## Abstract

Antibody-mediated rejection (AMR) is the leading cause of graft failure. While donor-specific antibodies (DSA) are associated with a higher risk of AMR, not all patients with DSA develop rejection suggesting that the characteristics of alloantibodies that determine their pathogenicity remain undefined. Using human leukocyte antigen (HLA)-A2-specific antibodies as a model, we applied systems serology tools to investigate qualitative features of immunoglobulin G (IgG) alloantibodies including Fc-glycosylation patterns and Fc**γ**R binding properties. The levels of afucosylation of anti-A2 antibodies were elevated in all seropositive patients and were significantly higher in AMR patients, suggesting potential cytotoxicity via Fc**γ**RIII-mediated mechanisms. Afucosylation of both glycoengineered monoclonal and naturally glycovariant polyclonal serum IgG specific to HLA-A2 exhibited potentiated binding to, slower dissociation from, and enhanced signaling through Fc**γ**RIII, a receptor widely expressed on innate effector cells. Collectively, these results suggest that afucosylated DSA may be a biomarker of AMR and could contribute to its pathogenesis.

**Graphical Abstract.**
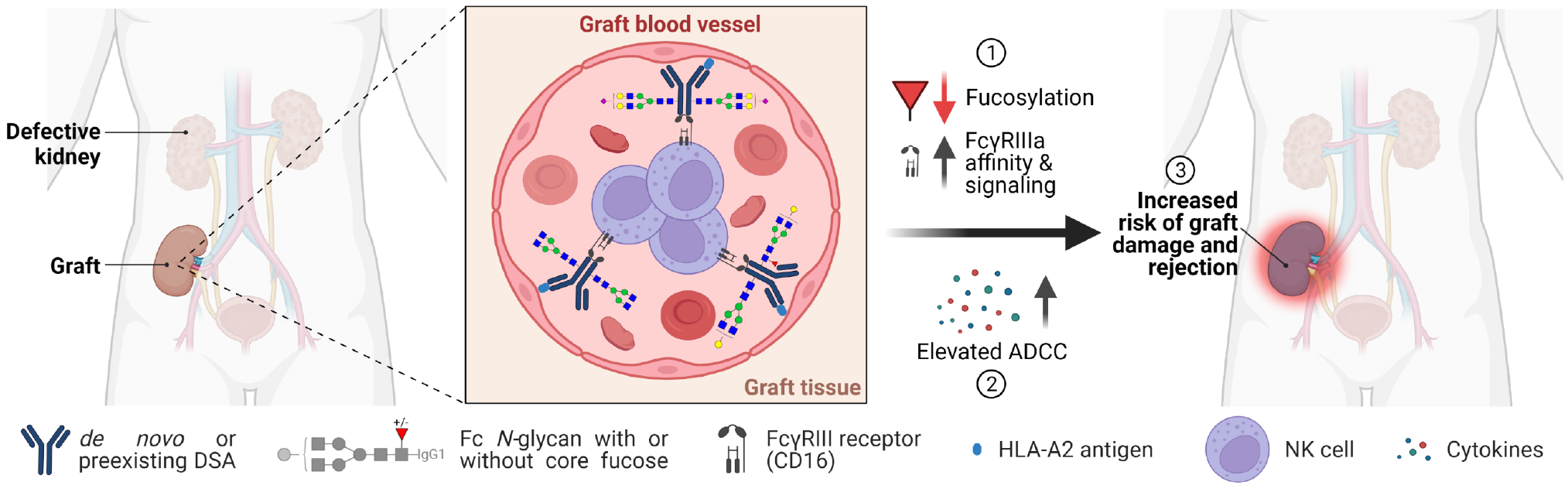
Potential influence of HLA-A2-specific IgG1 afucosylation, FcγRIIIa binding and activation on ADCC and graft rejection. Illustration created with https://BioRender.com.

## Introduction

Antibody-mediated rejection (AMR), encompassing allograft rejection caused primarily by antibodies directed against donor-specific human leukocyte antigen (HLA) molecules, is the leading cause of solid organ transplant rejection and long-term graft loss (1-5). AMR is characterized by histological manifestations of endothelial cell injury, mononuclear cell infiltration and complement-dependent tissue damage. Additionally, the presence of circulating donor-specific antibodies (DSA) poses challenges for transplant recipients by limiting access to available organs, prolonging wait time, and in some cases, excluding the candidates from a possible transplantation altogether (6-8). Solid phase assays using purified HLA antigens (Luminex® single bead antigen assays) have significantly improved stratification and categorization of transplant candidates, because they can detect very low levels of DSAs due to their high sensitivity. While this technique has been used to update organ allocation and desensitization protocols, it has led to minimal benefits to improvement in rejection treatment, as the presence and levels of DSA are not a reliable predictor of transplant outcome (9-13). While graft survival is clearly poorer for individuals sensitized against donor organ antigens (1, 2, 14-17), several studies have shown that not all DSAs carry the same risk of allograft rejection, as they have been associated with a wide spectrum of effects ranging from a complete absence of graft injury to the most severe form of AMR (18). Similarly, appearance of de novo DSA implies the risk of graft deterioration but provides little to no information on their actual pathogenic activities (3, 19-22).

Collectively, these observations point to the importance of factors other than the magnitude of the DSA response as important drivers of pathology. Clinical practice is challenged by the lack of strong relationships between antibody characteristics and patient outcomes. Given the profound disproportion between available organs and the number of patients waiting for a transplant, the US Food and Drug Administration (FDA) recently identified AMR and desensitization as two important areas in transplantation for which no drugs have been specifically approved (1, 2, 23-25). Techniques which better define DSAs properties may be required to first advance mechanistic understanding of AMR and secondly to improve transplant recipient outcomes.

To this end, the antibody effector functions that may be responsible for AMR are influenced not solely by titer, but by affinity, antigen availability and epitope, and antibody isotype, subclass, and glycosylation (12, 26-31). In other disease settings, systematic tools for surveillance of this spectrum of serum antibody features and their associated effector functions have identified reliable associations and begun to support robust predictions of disease outcomes (32-39). While prior research on the role of antibodies in transplant rejection was predominantly focused on estimation and retrospective correlation of titer to better predict transplant outcomes (16, 17, 40-42), recent studies have begun to interrogate other important factors governing antibody functionality, such as subclass distribution and complement fixing ability (12, 27, 28, 43-51), and the evolution of these features with the progression of disease (13, 48, 52-57).

Here, we report on the development and application of novel assays to characterize phenotypic and functional aspects of HLA-specific antibodies. Among a group of individuals with antibodies against HLA-A2, we observe altered Fc glycosylation profiles as compared to total serum IgG – most notably, enrichment of afucosylated IgG1 antibodies, which are widely associated with potentiated antibody-dependent cellular cytotoxicity (ADCC) (58-63). For both glycoengineered monoclonal as well as polyclonal, human serum-derived HLA-A2-specific antibodies, binding to, and signaling via Fc**γ**RIIIa, the receptor expressed on NK and other innate immune cells and responsible for mediating ADCC, were negatively associated with fucosylation. Collectively, this work establishes the potential importance of DSA Fc glycosylation in influencing the ADCC activity of DSA, and in turn, their ability to contribute to graft rejection.

## Methods

### Sample selection and study cohort

Clinical sample collection and analysis were approved by the Ethics Review Board of the Hôpital Erasme, Brussels, Belgium. Written informed consent was obtained from study participants. Clinical characteristics were collected from the patient’s electronic medical records (Transkid-RedCap). **(Supplemental Tables 1, 2)**. Patients with detectable anti-HLA-A2 alloantibodies were included in the study. Patients without detectable anti-A2 antibodies were included as controls. Anti-HLA-A2 antibodies were detected using clinical Luminex® single antigen bead assay according to the manufacturer’s instructions (Immucor^®^ Lifecodes). Following incubation with serum, Mean Fluorescence Intensity (MFI) of HLA antigen-coated beads was measured with a fluoroanalyser using xPonent software for data acquisition and Match It! Software (Immucor®Lifecodes) for data analysis. Positivity thresholds were defined according to the manufacturer’s instructions. For individuals with renal transplant, AMR was diagnosed based on clinical parameters (serum creatinine and proteinuria) and renal transplant biopsy, when contributive. AMR histological characteristics were glomerulitis, peritubular capillaritis, microvascular inflammation and C4d immunostaining positivity.

### Cloning and expression of recombinant HLA-A2 mAbs

Variable domain gene sequences of the heavy and the light chain (V_H_ and V_L_) of mouse HLA-A2 hybridoma cells (ATCC, HB-82 BB7.2) were defined to support recombinant production of a panel of human subclass-switched chimeric antibodies. Briefly, mRNA was isolated from hybridoma cells using the RNeasy kit (Qiagen, Cat No./ID: 74104), and cDNA generated using the VRSO cDNA kit (ThermoFisher, AB1453A). This cDNA was then amplified using degenerate primers (65) to selectively amplify V_H_ and V_L_ regions, which were sequenced and cross-referenced and annotated using BLAST and IMGT-based tools. Verified V_H_ and V_L_ sequences were used to design gene blocks (Twist Biosciences) that contained murine V_H_, V_L_, C_L_ and C_H_1 domains paired with human hinge and C_H_2 and C_H_3 Fc domains for each IgG subtype (**Supplemental Table 3**). These gene blocks were cloned by overlap extension into the pCMV expression plasmid.

Chimeric antibodies were transiently expressed via heavy and light chain plasmid co-transfection in HEK-expi293 cells, and purified using Protein A (IgG1, IgG2, and IgG4) or Protein G (IgG3) chromatography as previously reported (66, 67). Afucosylated IgG1 and IgG3 were produced by adding 0.15 mM of 2-fluorofucose (2FF) substrate in the growth medium, as described (67).

### IgG subclass and titer measurements

A custom multiplex assay was performed as previously described (68, 69) in order to define the total levels and subclass profiles of HLA-A2-, HLA-A1- (NIH Tetramer Facility, HLA-A*02:01 complexed with either CLGGLLTMV peptide from Epstein Bar Virus membrane protein or GLCTLVAML peptide from Epstein Bar Virus mRNA export factor ICP27, HLA-A*01:01 complexed with VTEHDTLLY from cytomegalovirus pp50), and HSV-gD- (Immune Technology IT-005-055p) specific antibodies. Briefly, antigen-coupled microspheres were diluted in Assay Buffer (PBS + 0.1% BSA + 0.05% Tween20), and mAb or serum, followed by washing and detection with R-phycoerythrin (PE)-conjugated anti-Human IgG (Southern Biotech #9040-09), anti-mouse IgG (Biolegend #405307) or anti-human IgG1 (#9054-09), IgG2 (Southern Biotech #9070-09), IgG3 (Southern Biotech # 1100-09) and IgG4 (Southern Biotech #9200-09), respectively. Median fluorescent intensities (MFI) were acquired on a FlexMap 3D (Luminex).

### Affinity purification of HLA-A2-specific antibodies

HLA-A2-specific antibodies were purified from human sera using magnetic, antigen-conjugated beads **(Supplemental Table 4)**. Magnetic streptavidin coated beads (NEB, S1420S) were incubated with biotinylated HLA-A2. Briefly, 50 μL streptavidin beads were washed with wash buffer (0.5M NaCl, 20mM Tris-HCl (pH 7.5), 1mM EDTA) followed by incubation with 20 μg biotinylated HLA-A2 for 2 hours at room temperature or overnight at 4°C. After washing five times, beads were blocked using 200 μL heat inactivated FBS for 2 hours. For purification, 150 μL of beads were co-incubated with 50 μL of sera for 3 hours on a rotational mixer, followed by three washes using PBS-TBN (PBS-1X, 0.1% BSA, 0.02% Tween 20, 0.05% sodium azide, pH 7.4). HLA-A2-specific antibodies were eluted by resuspending beads in 50 μL of 1% formic acid (pH 2.9) and incubating on a rotational mixer for 10 min at room temperature. Supernatant was withdrawn and 20 μL of 0.5 M sodium phosphate dibasic was added to each tube to neutralize the pH. The resulting eluate was split into two parts and used for LC-MS-based IgG Fc glycosylation analysis as well as for enrichment confirmation analysis, as described above.

### Sample preparation for LC-MS based Fc glycosylation analysis

Affinity purified HLA-A2-specific IgG were subjected to tryptic digestion overnight at 37 °C using sequencing grade trypsin (overall 25 ng) (Promega Corporation, WI, Madison), after the addition of 7 μL 1x PBS to each sample. Prior to LC-MS analysis, tryptic HLA-A2-specific IgG glycopeptides were desalted by reverse-phase solid phase extraction, as described (70). Total IgG was captured from plasma/serum using protein G Sepharose Fast Flow 4 beads (GE Healthcare, Uppsala, Sweden), as previously described (71).

### LC-MS based Fc glycosylation analysis and data processing

Glycopeptides were separated and detected with an Ultimate 3000 high-performance liquid chromatography (HPLC) system (Thermo Fisher Scientific, Waltham, MA) coupled to a maXis quadrupole time-of-flight mass spectrometer (Bruker Daltonics, Billerica, MA), as described (72). Data processing was performed according to established procedures (73).

### FcγR binding assay

The Fc**γ**R binding profiles of polyclonal HLA-A2-specific antibodies were defined using the Fc Array assay described previously (74). Briefly, HLA-A2-coated microspheres were generated as described above for the multiplex assay, but serum antibodies were detected with Fc**γ**RIIIa tetramers formed by mixing biotinylated Fc**γ**RIIIa V158 (75) with a 1/4^th^ molar ratio of Streptavidin-RPE (Agilent, PJ31S-1).

### FcγRIIIa signaling assay

As a surrogate for the ADCC potential of the serum-derived and recombinant HLA-A2-specific antibodies, Fc**γ**RIIIa signaling was measured by using a Jurkat Lucia NFAT reporter cell line (InvivoGen, jktl-nfat-cd16) (76, 77) in which cross-linking of antigen-bound antibodies and Fc**γ**RIIIa leads to the secretion of luciferase into the cell culture supernatant. Levels of Lucia luciferase secreted can then be directly measured by bioluminescence. Briefly, a clear flat bottom, high binding 96 well plate (Corning, 3361) was coated with either 1 μg/ml NeutrAvidin (ThermoFisher Scientific, 31000) or 1μg/mL biotinylated HLA-A2 antigen via incubation at 4°C overnight. The plates were then washed with 1X phosphate-buffered Saline (PBS) plus 0.05% Tween 20 and blocked with 1x PBS plus 2.5% bovine serum albumin (BSA) for 1 hour at room temperature. HLA-A2 antigen coated plates were directly used, whereas the plates coated first with NeutrAvidin were washed and incubated with 2μg/mL biotinylated HLA-A2 antigen for 1 hour at room temperature. After washing, 200 μL of serum diluted 1:100 and 100,000 cells/well in growth medium lacking antibiotics were added and incubated at 37°C for 24 hours. Alternatively, for HLA-A2 mAbs, the plates were first incubated with mAb for 3 hours at room temperature, washed and then was incubated with cells. The following day, 25 μL of supernatant was drawn from each well and then transferred to an opaque, white, clear bottom 96-well plate (Thomas Scientific, 6916A05) to which 75 μL of freshly prepared QuantiLuc (InvivoGen, rep-qlc2) was added. Luminescent was measured immediately on a SpectraMax Paradigm plate reader (Molecular Devices) using 1s of integration time for collecting light. The reported values are the means of three kinetic readings collected at 0, 2.5 and 5 minutes. Treatment of reporter cells with cell stimulation cocktail (ThermoFisher Scientific, 00-4970-93) was used as a positive control, and an **α**4**β**7-specific mAb was used as a negative control. Baseline control wells contained assay medium instead of antibody sample.

### FcγR affinity measurements

High Precision Streptavidin 2.0 (SAX2) biosensor tips (Sartorius) were used to determine the kinetics of HLA-A2-specific IgG binding to human Fc**γ**RIIIa and Fc**γ**RI by biolayer interferometry (BLI) on an Octet (Forte Bio) instrument. Streptavidin (SA) biosensor tips were first loaded with HLA-A2 antigen, followed by HLA-A2 antibody, and finally dipped into Fc**γ**R to study the antibody-receptor kinetics. Biosensors were equilibrated with 0.05% Tween-PBS for 60s, loaded with 0.25 µg/mL biotinylated HLA-A2 antigen (in 0.05% Tween-PBS) until they reached the threshold of 1nm, and then dipped into 0.05% Tween-PBS to reach baseline for 60s. After that, the biosensors were loaded with 3µg/mL of recombinant HLA-A2 IgG (in 0.05% Tween-PBS) for 600s, dipped into 0.05% Tween-PBS to reach baseline for 60s, and then dipped into 5μM monomeric Fc**γ**RIIIa for 180s or 292 nM Fc**γ**RI for 300s for association. To improve signal to noise, serum IgG was purified using Melon gel (Thermo) according to the manufacturer’s instructions. Polyclonal serum IgG was loaded for 2400s. For Fc**γ**R dissociation rate (k_d_) determination, two SAX2 biosensor tips were used, one for the measurement and one as a reference to subtract background signal changes due to dissociation of other components, such as dissociation of HLA-A2-specific IgG from biotinylated HLA. Reference biosensors loaded with IgGs were dipped into buffer (0.05% Tween-PBS) instead of receptors. Finally, the sensors were dipped into 0.05% Tween-PBS for 180s and 300s for the dissociation of Fc**γ**RIIIa and Fc**γ**RI, respectively. Data analysis

### Data analysis

Statistical analysis was performed in GraphPad Prism version 9. Replicates and statistical tests used are reported in figure legends. A nonlinear regression using a one phase exponential decay model was used to fit the dissociation curve and calculate off-rate.

## Results

HLA-specific antibody responses were investigated in a cohort consisting of individuals with reported clinical HLA-A2 reactivity (n=32) as detected with a single bead antigen assay, and controls (n=18) with no HLA-A2 reactivity. Among anti-A2 sensitized patients, 28 had received an organ transplant, including 26 kidney transplants and 2 other organs, and 4 patients were on a transplant waiting list (**Supplemental Table 1**). A2 sensitization was related to previous or current transplantation in 13 patients (41%), pregnancy in 6, blood transfusion in 1 and left ventricular assistance device in 1, whereas the sensitizing event was unknown in 4 patients. AMR was diagnosed in 13/28 (46%) transplanted patients (**Supplemental Table 1**) and in 7/13 (54%) DSA-positive patients (A2 sensitization in a patient transplanted with an HLA-A2 graft) **(Supplemental Table 2)**. Among controls, 9 patients had no detectable anti-HLA antibodies and 9 had detectable antibodies against non-HLA-A2 antigens. Seventeen controls had received an organ transplant and one was on a transplant waiting list.

### IgG subclass distribution of HLA-specific antibodies

We first sought to characterize the subclasses of HLA-specific IgGs in HLA-A2 sensitized and control subjects, using research grade single-antigen bead immunoassays. Briefly, single antigens (intact HLA-A02:01 or HLA-A01:01 monomers) were captured on microspheres, and antigen binding of sera samples were measured by multiplex assay. In contrast to the HIV-specific antibody VRC01 (negative control), the HLA-A2-specific monoclonal antibody BB7.2 (positive control), which was evaluated in each human IgG subclass, showed specific binding to HLA-A2 (**Figure 1A**), and appropriate staining by IgG subclass detection reagents (**Supplemental Figure 1**). In contrast to the negative control mAb, both pooled human serum immunoglobulin (IVIG) and serum from individual control subjects often showed signal considerably above background. While this profile may be consistent with the widespread prevalence of anti-HLA antibodies among healthy individuals (78, 79), and responses among subjects clinically defined as HLA-A2 seropositive were significantly elevated as compared to controls, there was a considerable overlap in distributions. Nonetheless, there was a distinct difference observed in the subclass distribution between the two groups, with the HLA-A2 seropositive group generally exhibiting a wide range of total HLA-A2 antibodies as compared to the control samples, which tended to show lower levels **(Figure 1A)**. The wide distribution of HLA-A2 specific IgG signal was consistent with reported values from clinical testing (**Supplemental Figure 2**), with some samples exhibiting moderate to very low HLA-A2 reactivity. HLA-A2-reactive antibodies predominantly belonged to the IgG1 and IgG2 subclasses, but a few subjects also exhibited higher levels of IgG3 and IgG4; these individuals typically had high levels of total HLA-A2-specific IgG.

**Figure 1.**
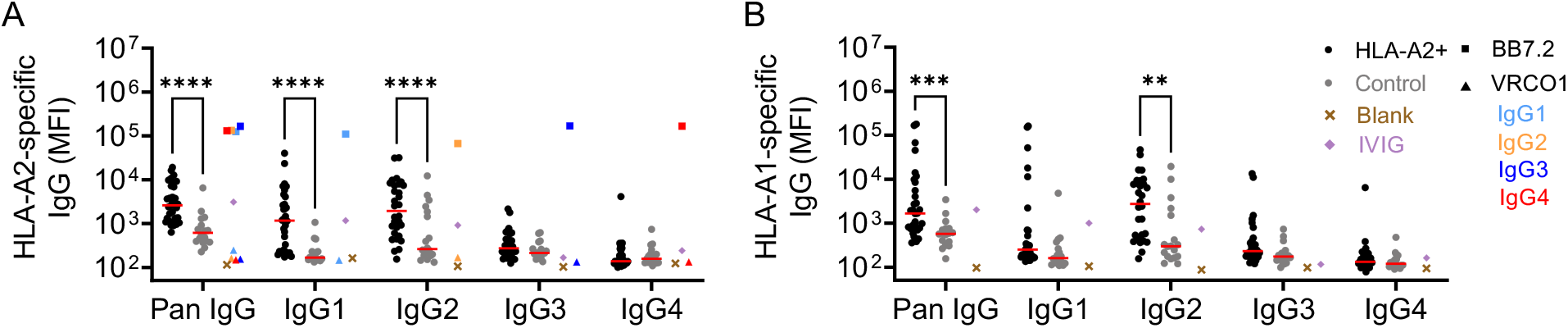
Subclass distribution of HLA specific antibodies. Characterization of HLA-specific antibodies binding to HLA-A2 **(A)** and HLA-A1 **(B)** antigens across IgG subclasses in HLA-A2 positive (black circles; n=30-32) and control (gray circles; n=18) individuals. HLA-A2-specific BB7.2 (square) and HIV-specific VRCO1 (triangle) mAb subclass controls for HLA-A2 reactivity are shown for each subclass: IgG1 (light blue), IgG2 (orange), IgG3 (dark blue) and IgG4 (red). Buffer only blank (cross) and pooled IVIG (diamond) are shown in brown and purple, respectively. Serum samples were tested at a 1:100 dilution. Data shown are representative of two technical replicates. Solid red lines indicate group median. Differences between groups were evaluated using Ordinary Two-way ANOVA adjusted for multiple comparisons using Bonferroni’s test (\*\**p*<0.01, \*\*\**p*<0.001, \*\*\*\**p* <0.0001, respectively).

A similar analysis of HLA-A1-specific antibody levels was performed. The HLA-A2 seropositive group also showed elevated levels of HLA-A1 antibodies as compared to the control group **(Figure 1B**), again, consistent with clinical single-antigen bead assay results. Interestingly, HLA-A1-specific antibodies predominantly belonged to the IgG2 subclass, with some samples exhibiting higher levels of IgG1, IgG3, and IgG4 subclasses.

### Affinity-enrichment yields HLA-A2-specific antibodies

A microscale purification method was successfully developed and employed for the purification of a control murine HLA-A2 mAb spiked into IVIG (**Supplemental Figures 3, 4**). Purified antibodies showed elevated reactivity to HLA-A2 antigen, but not to HLA-A1 or to an HSV antigen, which were used as negative controls, confirming the purity of the antibodies. Having vetted the purification process, HLA-A2-specific antibodies were then purified from serum of HLA-A2 antibody positive patients. The purified antibody fractions showed elevated reactivity to HLA-A2, but not to HSV. Despite generally being thought not to possess shared epitopes, the HLA-A2-enriched fraction showed some cross-reactivity with HLA-A1 for a few subjects who were generally strongly positive for both HLA-A1 and HLA-A2 antibodies (based on clinical single antigen bead assay) **(Supplemental Figure 5)**.

### HLA-A2 specific antibodies exhibit variable degree of afucosylation

Having confirmed the largely selective enrichment of HLA-A2-reactive antibodies, next we analyzed Fc glycosylation of both HLA-A2-specific and total serum IgG among HLA-A2 positive individuals. Fc glycosylation analysis revealed differences both on the level of individual glycoforms **(Figure 2A, B, C)**, and overall glycosylation traits including levels of fucosylation, bisection, galactosylation and sialyation (**Figure 2D**) between HLA-A2-specific and total serum IgG1. Of note, while HLA-A2-specific antibody Fc fucosylation of some individuals was comparable to that of total IgG1, others exhibited fucosylation levels reduced by 5-15%, or even 30% as compared to their total IgG1 counterpart **(Figure 2D)**. HLA-A2 specific IgG2/3 was characterized by higher bisection, galactosylation and sialylation than total serum IgG2/3, but no afucosylated IgG2/3 glycopeptides were detected **(Figure 2D)**. Consistent with multiplex assay data, antigen-specific IgG4 responses were generally too low to be robustly characterized by LC-MS. While IgG2 and IgG3 glycopeptides could not be distinguished by mass spectrometry due to their identical molecular mass, the high levels of antigen-specific IgG2 and low levels of IgG3 observed suggest that the antigen-specific IgG2/3 glycosylation signatures were dominated by IgG2.

**Figure 2.**
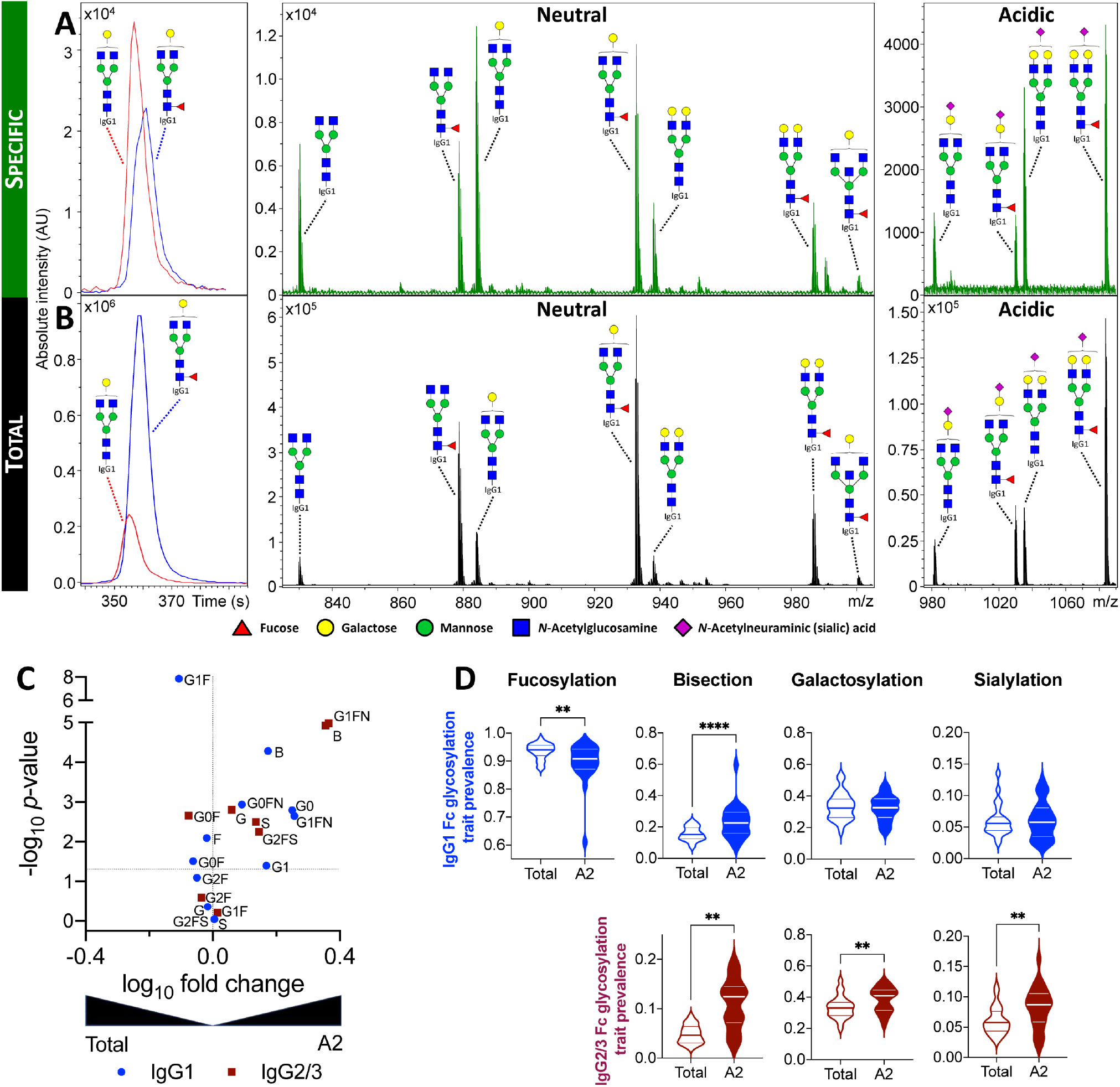
Fc glycosylation of HLA-A2-specific antibodies. Representative extracted ion chromatograms and mass spectra **(A, B)** illustrating the observed variability between HLA-A2-specific **(A)** and total **(B)** IgG1 Fc glycosylation patterns of the same patient. **C**. Volcano plot displaying the log_10_ fold change (x-axis) and -log_10_ *p*-value (y-axis) of individual IgG1 (blue circle) and IgG2/3 (maroon square) glycoforms between total and HLA-A2-specific IgG1 and IgG2/3, respectively. **D**. Violin plots showing the relative prevalence of glycans on bulk and HLA-A2 specific IgG1 (top) and IgG2/3 (bottom), respectively. Statistical analysis was performed using a paired two-tailed Student’s t-test (**: *p*<0.01, ***: *p*<0.001, ****: *p*<0.0001, respectively).

### Afucosylated HLA-A2-specific mAbs exhibit enhanced FcγRIIIa signaling

To study the Fc**γ**R signaling characteristics of HLA-A2-specific antibodies, we produced afucosylated BB7.2 IgG1 and IgG3 mAbs and assessed signaling driven by ligation of Fc**γ**RIII, a receptor typically expressed by NK cells, in a reporter cell line (77) as a surrogate for ADCC activity. IgG1 and IgG3 isotypes of BB7.2 were found to show Fc**γ**RIIIa ligation and signaling activity, whilst neither the control IgG1 subclass nor IgG2 and IgG4 subclasses of BB7.2 showed activity above baseline (**Figure 3A**). As expected, afucosylated IgG1 and IgG3 demonstrated considerably enhanced Fc**γ**RIII signaling activity as compared to their non-glycoengineered counterparts.

**Figure 3.**
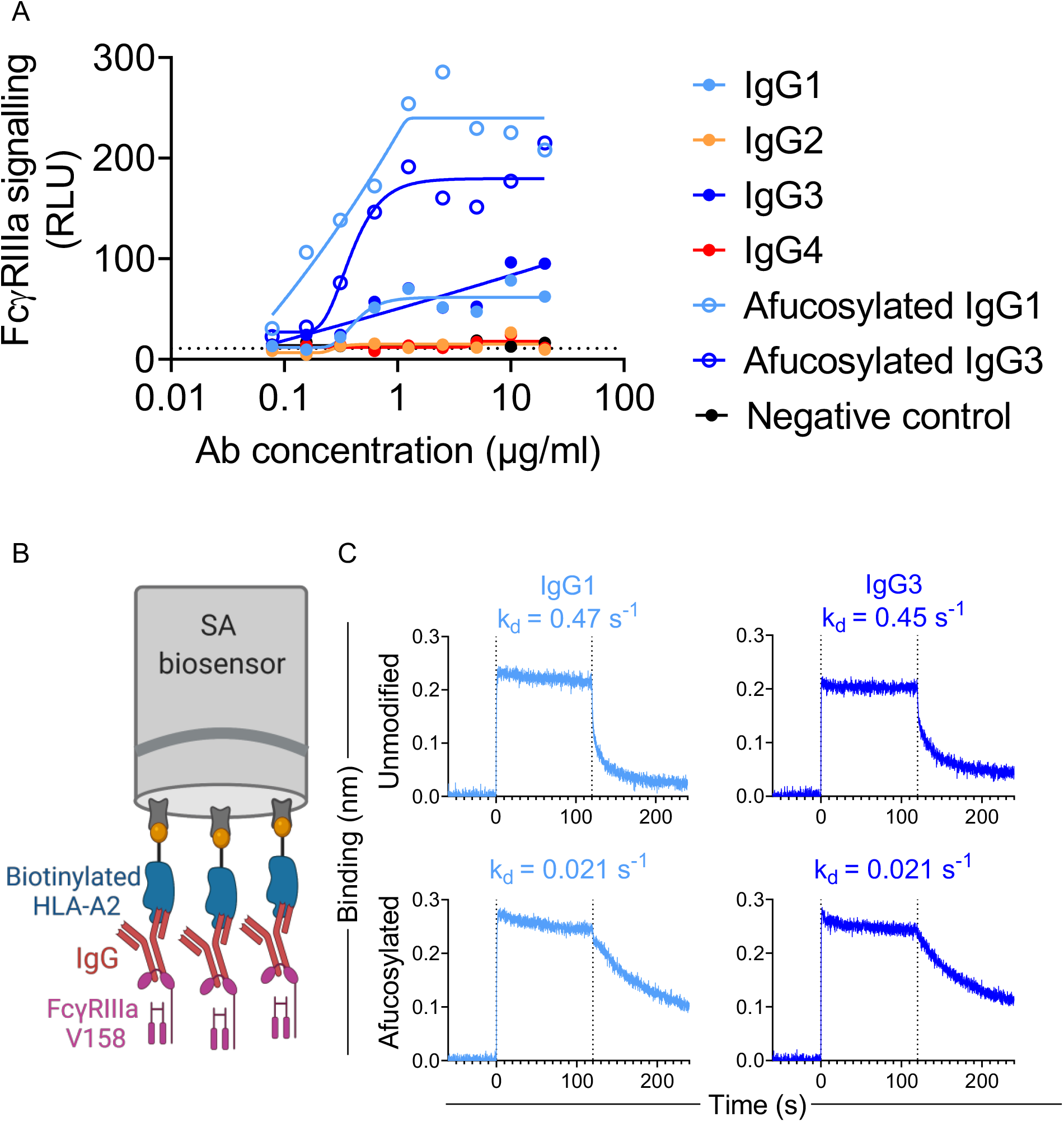
Impact of HLA-A2 specific mAb fucosylation on FcγRIIIa binding and signaling per subclass. **A**. Fc**γ**RIIIa signaling in a reporter cell line assay with unmodified and afucosylated HLA-A2 mAbs of varying subclasses. **B**. Schematic figure of the BLI experiment to define Fc**γ**RIIIa off-rates from HLA-A2-specific antibodies. Illustration created with http://BioRender.com. **C**. Fc**γ**RIIIa V158 association with and dissociation from unmodified and afucosylated HLA-A2 mAbs. Dissociation rate (k_d_) values are shown in inset.

### Enhanced FcγRIII signaling is associated with slow antibody dissociation

To address the mechanism of improved Fc**γ**RIIIa signaling activity, we next sought to characterize the interaction between HLA-A2-specific antibodies and Fc**γ**RIIIa using Biolayer Interferometry (BLI) (**Figure 3B, Supplemental Figure 6**), a label free technique commonly used to study the affinity and kinetics of protein-protein interactions. Antibodies bound to HLA-A2 antigen were allowed to bind (t = 0 to 120s) and then dissociate (t = 120-240s) from Fc**γ**RIIIa V158. All mAbs exhibited the fast on-rate typical of Fc**γ**R and showed similar levels of Fc**γ**RIIIa binding signal (**Figure 3C**). In contrast, dissociation rates (k_d_) varied by more than an order of magnitude between fucosylated and afucosylated mAbs **(Figure 3C)**. We found that afucosylated IgG1 and IgG3 had slower dissociation rates (mean k_d_ = 0.021 s^-1^) as compared to unmodified forms (mean k_d_ = 0.46 s^-1^). This difference in dissociation rate was specific for Fc**γ**RIIIa, as fucosylation did not influence Fc**γ**RI binding **(Supplemental Figure 7)**, as expected (63).

### Serum HLA-A2 specific antibody fucosylation associates with FcγRIIIa dissociation and ligation

Since afucosylated HLA-A2-specific mAbs had slower dissociation from Fc**γ**RIIIa as compared to their unmodified counterparts, we next studied the dissociation profiles of polyclonal HLA-A2 specific antibodies in human serum and found Fc**γ**RIIIa dissociation rate to be positively associated with fucosylation (**Figure 4A**). The variable off-rates observed among differentially fucosylated serum-derived HLA-A2-specific antibodies suggested that Fc**γ**RIIIa ligation and signaling profiles of these HLA-A2 specific antibodies might also vary. To test this possibility, HLA-A2 seropositive subjects were split into tertiles (high, medium, and low) based on their HLA-A2-specific antibody fucosylation levels **(Figure 4B)**. A multiplex assay was conducted, in which HLA-A2-specific antibodies were evaluated for their ability to bind Fc**γ**RIIIa tetramers (74). Although there was no significant difference in Fc**γ**RIIIa binding between high, medium, and low fucose tertiles, HLA-A2 specific antibodies in each of these three categories had higher Fc**γ**RIIIa binding as compared to negative controls, as exemplified by the observed negative correlations (**Figure 4B**). In line with observations on variably fucosylated mAbs, these results demonstrate that human serum-derived HLA-A2-specific antibodies with low fucosylation exhibit improved Fc**γ**RIIIa binding.

**Figure 4.**
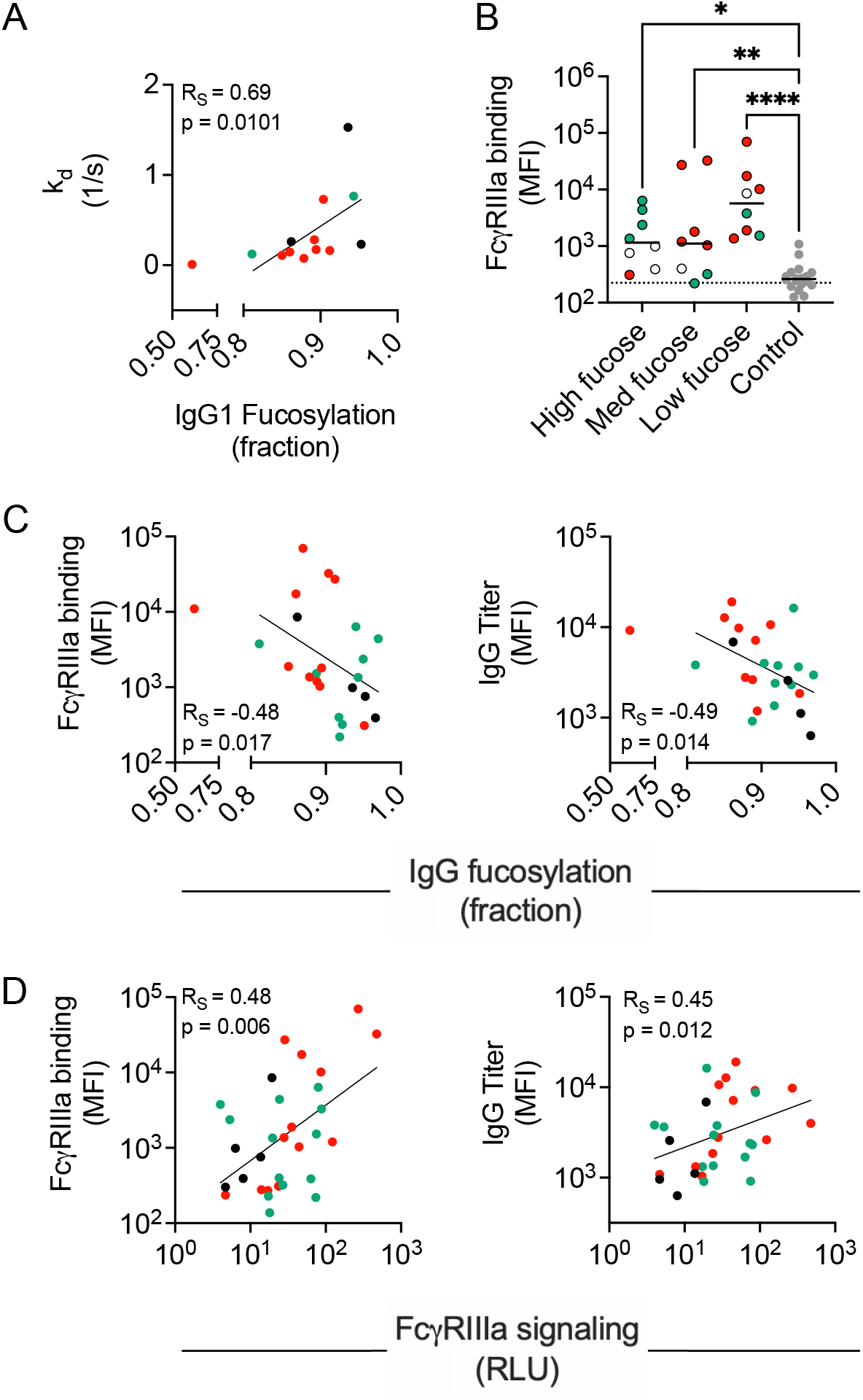
Associations of serum derived HLA-A2 specific antibody fucosylation with FcγRIIIa binding and signaling. **A**. Spearman’s correlation (R_S_) between IgG1 fucosylation and Fc**γ**RIIIa dissociation rate (n=13). **B**. Fc**γ**RIIIa binding characterization in high (n=8), medium (n=8), low (n=8) fucose samples and negative controls (n=18). Serum samples were tested at a 1:500 serum dilution. Statistical analysis was performed using Ordinary one-way ANOVA adjusted for multiple comparisons using Tukey’s test. Solid lines indicate group median. Data shown is representative of two technical replicates. **C-D**. Spearman’s correlations between IgG1 fucosylation (n=24) **(C)**, and Fc**γ**RIIIa signaling (n=31) **(D)** with Fc**γR**IIIa binding (left) and IgG titer (right). Patients with AMR (red), patients without AMR (green), and patients with no AMR information (black) are indicated in color.

### Serum HLA-A2 specific antibody fucosylation associates with functional activity

To understand how antibody fucosylation level was related to Fc**γ**RIIIa signaling, a limited correlation analysis was performed. Both HLA-A2-specific antibody titers and the Fc**γ**RIIIa ligation activity determined by multiplex assay were negatively associated with fucosylation **(Figure 4C)**, and positively associated with Fc**γ**RIIIa signaling **(Figure 4D)**. Despite the low signaling activity observed across experimental replicates in the reporter cell line assay **(Supplemental Figure 8A)**, relationships between Fc**γ**RIIIa signaling measured were consistently correlated to polyclonal HLA-A2-specific antibody titer and Fc**γ**RIIIa binding **(Supplemental Figure 8B)**.

### HLA-A2-specific antibody afucosylation was correlated with clinical AMR

To explore how HLA-A2 specific antibody characteristics relate to transplantation outcomes, patient data was analyzed based on AMR status. Subjects with AMR tended to have high HLA-A2 antibody titer and low HLA-A2-specific IgG1 fucosylation, thereby showing high Fc**γ**RIIIa binding, high signaling activity, and slower dissociation from Fc**γ**RIIIa **(Figure 4)**. As a more direct test, we analyzed HLA-A2 specific antibody features between subjects with and without AMR. Whether or not HLA-A2 represented a DSA, individuals with AMR showed significantly lower levels of HLA-A2-specific IgG1 fucosylation as compared to the individuals who did not have clinically defined AMR **(Figure 5A)**, suggesting that a low level of HLA-A2-specific antibody fucosylation may be a marker of AMR in kidney transplant patients. Other features, including HLA-A2-specific antibody titer, did not show such differences according to AMR status **(Supplemental Figure 9)**.

**Figure 5.**
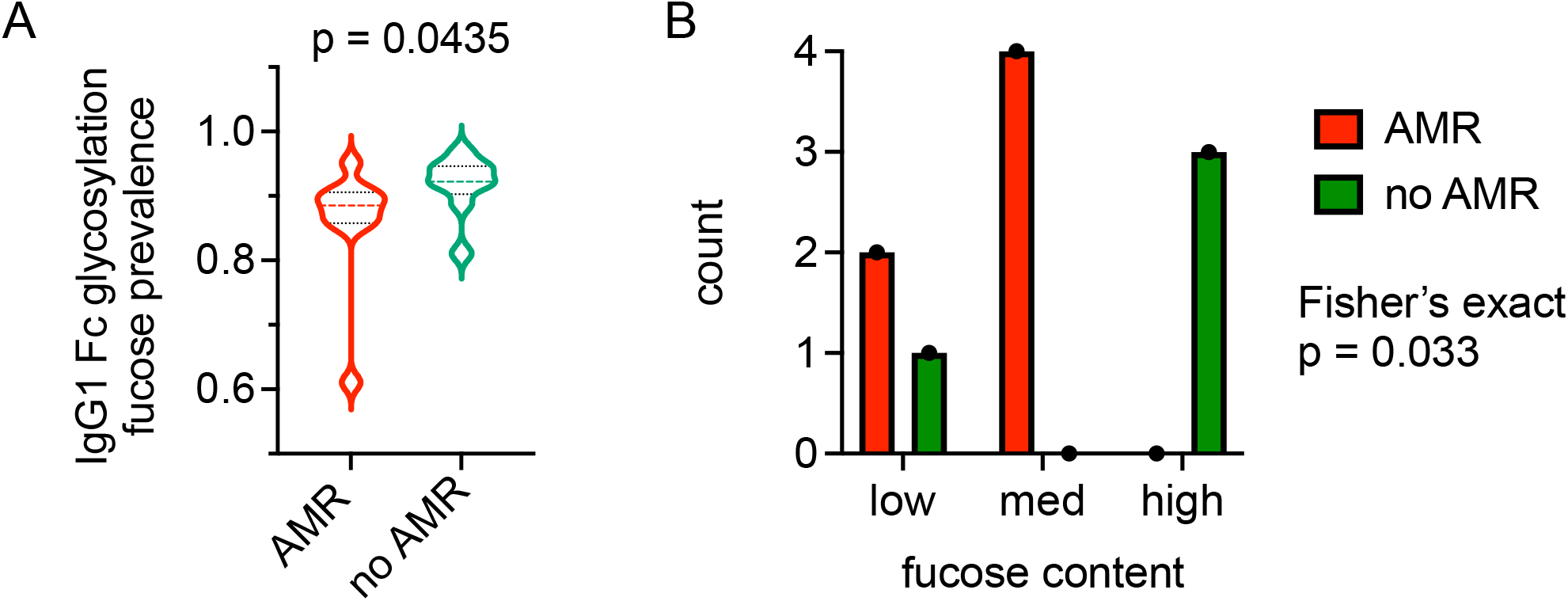
HLA-A2-specific IgG1 afucosylation is associated with AMR. **A**. Violin plot showing relative prevalence of fucose on HLA-A2-specific IgG1 in individuals with AMR (n=10) and without AMR (n=9). A Mann-Whitney U test was used to compare the two groups. **B**. Number of subjects in which HLA-A2-specific IgG1 comprise a DSA plotted by fucose content as tertiles (low, medium, high) and AMR status. Statistical significance defined by Fisher’s exact test.

In order to further examine the correlation between afucosylation of DSA and clinical AMR outcomes we analyzed subjects who had anti-HLA-A2 as a DSA. Patients who developed AMR had “low” and “moderate” fucose content of HLA-A2-specific DSA, whereas patients who did not develop AMR had “high” fucose content of HLA-A2-specific DSA. (**Figure 5B**). Collectively, this data suggests that the phenotypic variability among anti-HLA-A2 antibody responses may have an important influence on organ transplant outcomes.

## Discussion

The last two decades have witnessed a considerable increase in understanding immunological processes related to organ rejection, which has contributed to reduction in the incidence of acute rejection and improvement in short-term graft survival (23, 80-82). Despite the advances in immunosuppression strategies, HLA matching, and better management of acute rejection, the prognosis for long-term graft outcome remains one of the biggest challenges in the field of organ transplantation (2). Given the well-established importance of qualitative aspects of the antibody response in other settings, consideration of antibody subclass, Fc glycosylation, and Fc**γ**R binding in the context of renal transplant outcomes will define the importance of these features in mediating functions that escalate the risk of organ rejection (2). Discovery of robust relationships between features of DSA and transplant outcomes has the potential to drive revision of clinical strategies of organ allocation as well as development of novel interventions.

To date, relatively few studies have looked at DSA subclass and Fc glycosylation in kidney transplant recipients. Challenges in DSA purification from plasma/serum appear to have posed a major hurdle to its glycosylation analysis, resulting in a focus on characterization of total serum IgG glycosylation (83-85). In this study, we employed a microscale purification method to purify HLA-A2-specific antibodies from patient sera and set out to evaluate IgG subclass and Fc glycosylation profiles thereof. Both of these features regulate Fc-Fc**γ**R interactions, which in turn potentially relate to pathogenicity and graft rejection.

Beyond the complexity of the many processes associated with allograft injury and rejection, phenotypic changes in DSA over time have also been observed (43), complicating the understanding of the pathophysiology of rejection. What has become clear is that DSA titers, subclass profiles or C1q binding activity alone are insufficient to accurately predict the course of AMR, and the field remains in need of reliable prognostic biomarkers to guide treatment, optimally allocate organs, and reduce the risk of rejection in DSA-sensitized recipients.

Though not strongly associated with outcomes of organ rejection, IgG subclass is known to strongly modify the ability of antibodies to drive complement deposition and recruit the innate immune effector cells that mediate clearance of opsonized particles (5, 27, 29, 43, 86). HLA-A2-specific responses in clinically seropositive subjects were predominantly IgG1 and IgG2. High levels of IgG1 antibodies suggest the possibility of elevated Fc mediated effector functions, such as phagocytosis, ADCC and complement deposition (63), which might lead to severe graft injury and transplant rejection. The prevalence of HLA-A1-specific IgG2 antibodies in this population suggests that the IgG subclass profile can vary from one HLA antigen to another even within a given individual, highlighting the potential importance of integrating profiles across diverse specificities and characterizing features of anti-HLA antibodies beyond titer.

Antibody effector functions are highly affected by the composition of the *N*-glycan present at the conserved *N*-glycosylation site in the Fc region (60, 61, 87-89). Hitherto, it has been repeatedly shown that disease-specific antibodies can exhibit skewed glycosylation profiles, that in turn associate with disease prognosis and outcome (73, 90-93). Historically, one of the key limitations of glyco-profiling such antibodies is their low serum prevalence and high sample requirement. In order to facilitate Fc glycosylation analysis of low abundant HLA-A2 specific antibodies, an antigen-specific antibody purification approach was developed for reliable, sensitive, and specific capturing of HLA-A2 specific antibodies from reactive sera. This platform was leveraged to support analysis of HLA-A2-specific IgG Fc glycosylation profiles. As compared to the global serum IgG profiles, we observed variable degree of HLA-A2 specific IgG1s afucosylation. This feature was associated with improved Fc**γ**RIIIa binding as afucosylation has been described to lead to elevated Fc**γ**RIII binding (58, 63, 83-86), we hypothesized that low HLA-A2-specific IgG1 fucosylation could cause elevated DSA cytotoxicity via enhanced ADCC. Indeed, when tested in the context of a subclass-switched recombinant antibody specific for HLA-A2, both subclass and fucosylation had a strong impact on Fc**γ**RIIIa affinity and signaling. Specifically, afucosylated mAbs had slower dissociation from the receptor, which could provide sufficient time for the receptors to cross-link and activate downstream signaling pathways and modulate ADCC. This relationship was also observed among polyclonal HLA-A2-specific antibodies purified from patient sera, and suggests that afucosylated antibodies to HLA-A2 can exhibit elevated ADCC activity, as shown in other disease settings (70, 92, 94-96), and raises the possibility that DSA glycosylation may provide prognostic value in predicting risk of AMR. Indeed, individuals who had AMR exhibited low levels of HLA-A2 specific IgG1 fucosylation as compared to the individuals who did not have AMR, suggesting that low HLA-A2 IgG1 fucosylation may be a marker of AMR risk in kidney transplant patients. Despite small cohort size, among individuals in whom HLA-A2 represented a DSA, low HLA-A2 specific IgG1 antibody fucosylation was associated with AMR.

While this work suggests the potential role of IgG Fc fucosylation and its influence on DSA pathogenicity, it is likely that more systematic serology data, considering antibody titer, subclass, glycans, epitope specificity, antigen-specificity, and affinity, linked to clinical metadata and insights into host- and graft-specific genetic and phenotypic factors will be needed for comprehensive understanding. The experimental tools explored here will need to be broadened to cover the diversity of DSA specificities that contribute to clinical outcomes. Similarly, analytical approaches (97-99) that integrate across features of the humoral response, and that can robustly address correlations among features, such as those observed here between titer and afucosylation, may contribute to discovery of mechanisms underlying graft injury and AMR. To this end, the subclass-switched and glycoengineered mAbs described herein may support dissection of the mechanistic role of these features in AMR studies in animal models.

Limitations of this study include its focus on a single antigen-specificity and antibody function, limited outcome data and reliance on a relatively small group of HLA-A2 seropositive subjects. The lack of a clear boundary between seropositive and seronegative subjects based on clinical testing suggests that positivity determinations may be quite sensitive to different reagents, tests, and thresholds, as well as to differences in antigen density and loaded antigens between single antigen beads and research microspheres applied by different clinical groups. A reporter cell line and recombinant HLA-A2 antigen, as opposed to primary effector and HLA-A2-expressing target cells were used to evaluate Fc**γ**RIIIa signaling as a proxy for cytotoxicity, degranulation, and release of other inflammatory mediators that may be relevant to graft damage. Antibody clonality, fine epitope specificity, and affinity for HLA-A2 were not assessed. Despite extensive condition testing, signals in the reporter cell line assay were low for both mAbs and polyclonal sera samples, and despite their correlations with other activities, were not always elevated in comparison to the control sera. Similarly, while signals in the in-house multiplex assay were correlated with clinical test results, agreement was less good than we have observed in other multiplexed assays, and distributions overlapped with those observed among control sera, perhaps in association with the rate of seropositivity in healthy populations (78, 79). While similar yields were observed to result from affinity purification against HLA-A2 with each of two different peptides in a pilot experiment, peptide occupancy and identity may also impact results of the assays used. Lastly, while a direct association has been made between afucosylation of DSA and the occurrence of AMR, HLA-A2-specific antibodies were first detected in most of the patients post-rejection, suggesting that afucosylation of DSA may have occurred as a result of AMR. Future studies will benefit from larger and longitudinal cohorts that include samples collected pre-AMR, that are more homogenous in relevant clinical characteristics, and designs that are targeted to address relationships with transplant outcomes.

Overall, despite considerable clinical heterogeneity of the study population, HLA-A2-specific IgG fucosylation was associated with Fc**γ**RIIIa binding and function. Different HLA reactivities showed distinct subclass profiles, and the dominant IgG1 fraction of HLA-A2-specific IgG showed reduced fucosylation as compared to total serum IgG1. Along with antibody titer, reduced fucosylation was associated with elevated binding to, and slower dissociation from Fc**γ**RIIIa. Afucosylation of HLA-A2-specific antibodies was also associated with potentiated signaling through Fc**γ**RIIIa. HLA-A2-specific IgG1 Fc fucose content was associated with AMR status among the subset of subjects for whom this specificity represented a DSA as well among those where it did not, leaving important questions unresolved about the importance of this antibody feature in mediating AMR. Further and prospective studies will be needed to confirm the role of afucosylation in the pathophysiology of AMR, and continued customization and broader deployment of these and other systems serology approaches has the potential to shed light on the clinical impact of qualitative antibody features and activities on graft damage.

## Data Availability

Data produced in the present study are available upon reasonable request to the authors.

## Author contributions

**P**.**B**.: writing - original draft, investigation, data curation, visualization, methodology, validation; **S**.**S**.: writing - original draft, investigation, data curation, validation, visualization; **T**.**P**.: writing – review & editing, investigation, data curation, validation, visualization; **C**.**C**.:, **S**.**S**.:, **A**.**L**.**M**.: ; **N. de H**.: writing review & editing, supervision; **N**.**M**.: writing-review & editing, supervision; **L**.**V**.**R**.: ; **M**.**W**.: writing review & editing, funding acquisition, supervision; **A**.**M**.:, **M**.**E**.**A**.: conceptualization, supervision, funding acquisition, writing – review & editing.

## Acknowledgements

We want to thank Andreia Isabel Ferreira Verissimo and Emilie Shipman from bioMT-Institute for Biomolecular Targeting at Dartmouth. The authors also thank Carolien Koeleman for expert technical assistance. We thank all members of the HLA Laboratory, Laboratoire Hospitalier Universitaire de Bruxelles (LHUB), and Dr Sanâa El Korchi, Erasme Hospital ULB, Brussels, Belgium, for expert assistance in clinical data and sample management and in clinical sample testing. We thank the NIH Tetramer Core Facility (contract number 75N93020D00005) for providing HLA monomers.

## Data availability statement

The dataset generated for this study is available at (link pending).

## Abbreviations

ADCC: Antibody-dependent cellular cytotoxicity
AMR: Acute-mediated rejection
BLI: Biolayer Interferometry
DSA: Donor specific antibody
Fc: Fragment crystallizable
HLA: Human leukocyte antigen
IgG: Immunoglobulin G
IVIG: Intravenous immunoglobulin
G MFI: Mean Fluorescent Intensity
R: receptor

