## Supplemental Tables for "Afucosylation of HLA-specific IgG1 as a potential predictor of antibody pathogenicity in kidney transplantation"

| Supplemental Tables | |
|---|---|
| Supplemental Table 1 | Whole cohort characteristics |
| Supplemental Table 2 | DSA cohort characteristics |
| Supplemental Table 3 | HLA-A2-specific monoclonal antibody gene sequences |
| Supplemental Table 4 | Details of HLA molecules used for the purification of HLA-specific IgGs |

**Supplemental Table 1. Cohort characteristics.** The median and P25-P75 are shown, unless indicated otherwise.

| Characteristics | Anti-HLA-A2 sensitized group (n=32) | Control group (n=18) |
|---|---|---|
| **Patient Age - Years (P25-P75)** | 45.5 (28-57.5) | 48 (41-61) |
| **Patient Sex - n (%)**<br>Male<br>Female | <br>12 (37.5%)<br>20 (62.5) | <br>14 (77.7%)<br>4 (22.2%) |
| **Sensitizing event – n (%)**<br>Transplantation<br>Pregnancies<br>Blood derived product transfusions<br>Left ventricular assistance device<br>Unknown<br>No previous sensitizing event | <br>13 (40.6%)<br>6 (18.8%)<br>1 (3.1%)<br>1 (3.1%)<br>11 (34.4 %)<br>0 (0%) | <br>5 (27,8%)<br>2 (11,1%)<br>1 (5,6%)<br>0 (0%)<br>2 (11,1%)<br>8 (44,4%) |
| **Type of transplant among transplanted patients – n (%)**<br>KT<br>KPT<br>HT<br>PT<br>PLT<br>LKT<br>On transplant waiting list | 25 (78.1%)<br>1 (3.1%)<br>1 (3.1%)<br>1 (3.1%)<br>0 (0%)<br>0 (0%)<br>4 (12.5%) | 15 (83.3%)<br>0 (0%)<br>0 (0%)<br>0 (0%)<br>1 (5.5%)<br>1 (5.5%)<br>1 (5.5%) |
| **Donor sex – n (%)**<br>Male<br>Female<br>Unknown | <br>10 (35.7%)<br>16 (57.1 %)<br>2 (7.1%) | <br>13 (76,5%)<br>3 (17,6%)<br>1 (5,9%) |
| **Donor age among transplanted patients (P25-P75)** * | 44 (35-54) | 40.5 (31.75-50) |
| **Donor category among transplanted patients – n (%)**<br>Deceased<br>Living<br>Not reported | 22 (78.56%)<br>3 (10.7%)<br>3 (10.7%) | 15 (88.2%)<br>2 (11.8%)<br>0 (0%) |
| **Warm ischemia time among transplanted patients – minutes (P25-P75)** * | 32 (27.5-38.5) | 36.5 (24.5-43.25) |
| **Cold ischemia time among transplanted patients – hours (P25-P75)** * | 19 (13-21.5) | 20 (6.95-20.5) |
| **Recovery of graft function among transplanted patients – n (%)** | 20 (71.4%) | 14 (82,4%) |

| | | |
|---|---|---|
| Immediate<br>Delayed<br>Not reported | 1 (3.6%)<br>7 (25%) | 2 (11.8%)<br>1 (5,9%) |
| **HLA A-B-DR mismatches among transplanted patients – n (P25-P75)** * | 3 (2-3.5) | 3 (2-4) |
| **Induction immunosuppressive therapy among transplanted patients – n (%)**<br>No induction<br>Basiliximab<br>Thymoglobulin<br>OKT3<br>Unknown | <br><br>3 (10.7%)<br>6 (21.4%)<br>11 (39.3%)<br>1 (3.6%)<br>7 (25%) | <br><br>2 (11.8%)<br>10 (58.8%)<br>3 (17.6%)<br>1 (5.9%)<br>1 (5.9%) |
| **Maintenance immunosuppressive therapy among transplanted patients – n (%)**<br>CNI, MMF, mPDN<br>CNI, AZA, mPDN<br>Others<br>Not reported | <br><br>19 (67.8%)<br>3 (10.7%)<br>0 (0%)<br>6 (21.4%) | <br><br>14 (82.4%)<br>0 (0%)<br>2 (11.8%)<br>1 (5.9%) |
| **AMR – n (%)**<br>Unknown<br>No AMR<br>AMR<br>- Acute AMR<br>- Chronic AMR | <br>2 (7.1%)<br>13 (46.4%)<br>13 (46.4%)<br>3 (23.8%)<br>10 (76.9%) | <br>2 (11.8%)<br>12 (70.6%)<br>3 (17.6%)<br>2 (66.7%)<br>1 (33.3%) |
| **Anti-HLA-A2 fucosylation - n (%)**<br>Low<br>Medium<br>High<br>Undetermined | <br>8 (25%)<br>8 (25%)<br>8 (25%)<br>8 (25%) | Glycans not determined |
| **Time of anti-HLA-A2 antibodies detected among patients with AMR – n (%)**<br>Pre AMR<br>Post AMR | <br><br><br>2 (15.4%)<br>11 (84.6%) | |

* Data are given when available.

Abbreviations: KT: Kidney transplant, KPT: combined kidney pancreas transplant, HT: heart transplant, PT: pulmonary transplant, PLT: combined pulmonary liver transplant, LKT: combined liver kidney transplant.
CNI: calcineurin inhibitors; MMF: mofetil mycophenolate; mPDN methylprednisolone; AZA: Azathioprine; AMR: antibody mediated rejection.

**Supplemental Table 2. DSA cohort characteristics.** The median and P25-P75 are shown, unless indicated otherwise.

| **Characteristics** | **Anti-HLA-A2 DSA sub-group (n=13)** |
|---|---|
| **Patient age - Years (P25-P75)** | 30 (21-44) |
| **Sex - n (%)** | |
| Male | 9 (64.3%) |
| Female | 5 (35.7%) |
| **Type of transplant – n (%)** | |
| KT | 12 (92.3%) |
| KPT | 1 (7.69%) |
| PLT | 0 (0%) |
| LKT | 0 (0%) |
| On transplant waiting list | 0 (0%) |
| **Donor sex– n (%)** | |
| Male | 6 (46.1%) |
| Female | 7 (53.9%) |
| Unknown | 0 (0%) |
| **Donor age– years (P25-P75)** | 41 (22-54) |
| **Donor category – n (%)** | |
| Deceased | 11 (84.6%) |
| Living | 1 (7.7%) |
| Not reported | 1 (7.7%) |
| **Warm ischemia time – minutes (P25-P75)*** | 32 (28-33) |
| **Cold ischemia time – hours (P25-P75)*** | 20 (16-26.5) |
| **Recovery of graft function – n (%)** | |
| Immediate | 10 (76.9%) |
| Delayed | 0 (0%) |
| Not reported | 3 (23.1%) |
| **HLA A-B-DR mismatches – n (P25-P75)*** | 2 (2-3.5) |
| **Induction immunosuppressive therapy –n (%)** | |
| No induction | 1 (7.7%) |
| Basiliximab | 2 (15.4%) |
| Thymoglobulin | 5 (38.5%) |
| OKT3 | 1 (7.7%) |
| Not reported | 4 (30.7%) |

| | |
|---|---|
| **Maintenance immunosuppressive therapy – n (%)**<br>CNI, MMF, mPDN<br>CNI, AZA, mPDN<br>Others<br>Not reported | 6 (46.2%)<br>3 (23.1%)<br>0 (0%)<br>4 (30.8%) |
| **AMR – n (%)**<br>Not reported<br>No AMR<br>AMR<br>- Acute AMR<br>- Chronic AMR | <br>0 (0%)<br>6 (46.2%)<br>7 (53.8%)<br>3 (42.9%)<br>4 (57.1%) |
| **Time of anti-HLA-A2 antibodies detected among patients with AMR**<br>Post AMR | <br><br>7 (100%) |
| **Fucosylation profile among AMR positive patients - n (%)**<br>Low<br>Medium<br>High<br>No glycan information | <br><br>2 (28.57%)<br>4 (57.14%)<br>0 (0%)<br>1 (14.28%) |
| **Fucosylation profile among AMR negative patients – n (%)**<br>Low<br>Medium<br>High<br>No glycan information | <br><br>1 (16.66%)<br>0 (0%)<br>3 (50%)<br>2 (33.33%) |

* Data are given when available.

Abbreviations: KT: Kidney transplant, KPT: combined kidney pancreas transplant, HT: heart transplant, PT: pulmonary transplant, PLT: combined pulmonary liver transplant, LKT: combined liver kidney transplant.
CNI: calcineurin inhibitors; MMF: mofetil mycophenolate; mPDN methylprednisolone; AZA: Azathioprine; AMR: antibody mediated rejection.

**Supplemental Table 3. HLA-A2-specific monoclonal antibody gene sequences.**

| Gene sequence | |
|---|---|
| **light chain** | gatgttttgatgacccaaactccactctccctgcctgtcagtcttggagatcaagtctccatctcttgcagatctagtcaga<br>gcattgtacatagtaatggaaacacctatttagaatggtacctgcagaaaccaggccagtctccaaagctcctgatctac<br>aaagtttccaaccgattttctggggtcccagacaggttcagtggcagtggatcagggacagatttcacactcaagatca<br>gcagagtggaggctgaggatctgggagtttattactgctttcaaggttcacatgttcctcggacgttcggtggaggca<br>ccaagctggaaatcaaaCGGGCAGATGCTGCACCAACTGTATCCATCTTCCCACCATCCAG<br>TGAGCAGTTAACATCTGGAGGTGCCTCAGTCGTGTGCTTCTTGAACAACTTCTACCC<br>CAAAGACATCAATGTCAAGTGGAAGATTGATGGCAGTGAACGACAAAATGGCGTC<br>CTGAACAGTTGGACTGATCAGGACAGCAAAGACAGCACCTACAGCATGAGCAGCA<br>CCCTCACGTTGACCAAGGACGAGTATGAACGACATAACAGCTATACCTGTGAGGCC<br>ACTCACAAGACATCAACTTCACCCATTGTCAAGAGCTTCAACAGGAATGAGTGT |
| **IgG1 heavy chain** | caggtccagctgcagcagtctggacctgagctggtgaagcctggggcctcagtgaagatgtcctgcaaggcttctgg<br>ctacaccttcacaagctaccatatacagtgggtgaagcagaggcctggacagggacttgagtggattggatggattta<br>tcctggagatggtagtactcagtacaatgagaagttcaagggcaagaccacactgactgcagacaaatcctccagcac<br>agcctacatgttgctcagcagcctgacctctgaggactctgcgatctatttctgtgcaagggaggggacctactatgct<br>atggactactggggtcaaggaacctcagtcaccgtctcctcaGCTAAAACAACACCCCCATCAGTCTA<br>TCCACTGGCCCCTGGGTGTGGAGATACAACTGGTTCCTCTGTGACTCTGGGATGCC<br>TGGTCAAGGGCTACTTCCCTGAGTCAGTGACTGTGACTTGGAACTCTGGATCCCTGT<br>CCAGCAGTGTGCACACCTTCCCAGCTCTCCTGCAGTCTGGACTCTACACTATGAGC<br>AGCTCAGTGACTGTCCCCTCCAGCACCTGGCCAAGTCAGACCGTCACCTGCAGCGT<br>TGCTCACCCAGCCAGCAGCACCACGGTGGACAAAAAACTTGAGCCCAAATCTTGT<br>GACAAAACTCACACATGCCCACCGTGCCCAGCACCTGAACTCCTGGGGGGACCGT<br>CAGTCTTCCTCTTCCCCCCAAAACCCAAGGACACCCTCATGATCTCCCGGACCCCT<br>GAGGTCACATGCGTGGTGGTGGACGTGAGCCACGAAGACCCTGAGGTCAAGTTCA<br>ACTGGTACGTGGACGGCGTGGAGGTGCATAATGCCAAGACAAAGCCGCGGGAGG<br>AGCAGTACAACAGCACGTACCGTGTGGTCAGCGTCCTCACCGTCCTGCACCAGGA<br>CTGGCTGAATGGCAAGGAGTACAAGTGCAAGGTCTCCAACAAAGCCCTCCCAGCC<br>CCCATCGAGAAAACCATCTCCAAAGCCAAAGGGCAGCCCCGAGAACCACAGGTGT<br>ACACCCTGCCCCCATCCCGGGAGGAGATGACCAAGAACCAGGTCAGCCTGACCTG<br>CCTGGTCAAAGGCTTCTATCCCAGCGACATCGCCGTGGAGTGGGAGAGCAATGGG<br>CAGCCGGAGAACAACTACAAGACCACGCCTCCCGTGCTGGACTCCGACGGCTCCT<br>TCTTCCTCTACAGCAAGCTCACCGTGGACAAGAGCAGGTGGCAGCAGGGGAACGT<br>CTTCTCATGCTCCGTGATGCATGAGGCTCTGCACAACCACTACACGCAGAAGAGCC<br>TCTCCCTGTCTCCGGGTAAA |

| | |
|---|---|
| **IgG2 heavy chain** | caggtccagctgcagcagtctggacctgagctggtgaagcctggggcctcagtgaagatgtcctgcaaggcttctgg<br>ctacaccttcacaagctaccatatacagtgggtgaagcagaggcctggacagggacttgagtggattggatggattta<br>tcctggagatggtagtactcagtacaatgagaagttcaagggcaagaccacactgactgcagacaaatcctccagcac<br>agcctacatgttgctcagcagcctgacctctgaggactctgcgatctatttctgtgcaagggaggggacctactatgct<br>atggactactggggtcaaggaacctcagtcaccgtctcctcaGCTAAAACAACACCCCCATCAGTCTA<br>TCCACTGGCCCCTGGGTGTGGAGATACAACTGGTTCCTCTGTGACTCTGGGATGCC<br>TGGTCAAGGGCTACTTCCCTGAGTCAGTGACTGTGACTTGGAACTCTGGATCCCTGT<br>CCAGCAGTGTGCACACCTTCCCAGCTCTCCTGCAGTCTGGACTCTACACTATGAGC<br>AGCTCAGTGACTGTCCCCTCCAGCACCTGGCCAAGTCAGACCGTCACCTGCAGCGT<br>TGCTCACCCAGCCAGCAGCACCACGGTGGACAAAAAACTTGAGCGCAAATGTTGT<br>GTCGAGTGCCCACCGTGCCCAGCACCACCTGTGGCAGGACCGTCAGTCTTCCTCTT<br>CCCCCCAAAACCCAAGGACACCCTCATGATCTCCCGGACCCCTGAGGTCACGTGC<br>GTGGTGGTGGACGTGAGCCACGAAGACCCCGAGGTCCAGTTCAACTGGTACGTGG<br>ACGGCGTGGAGGTGCATAATGCCAAGACAAAGCCACGGGAGGAGCAGTTCAACA<br>GCACGTTCCGTGTGGTCAGCGTCCTCACCGTTGTGCACCAGGACTGGCTGAACGG<br>CAAGGAGTACAAGTGCAAGGTCTCCAACAAAGGCCTCCCAGCCCCCATCGAGAAA<br>ACCATCTCCAAAACCAAAGGGCAGCCCCGAGAACCACAGGTGTACACCCTGCCCC<br>CATCCCGGGAGGAGATGACCAAGAACCAGGTCAGCCTGACCTGCCTGGTCAAAGG<br>CTTCTACCCCAGCGACATCGCCGTGGAGTGGGAGAGCAATGGGCAGCCGGAGAA<br>CAACTACAAGACCACGCCTCCCATGCTGGACTCCGACGGCTCCTTCTTCCTCTACA<br>GCAAGCTCACCGTGGACAAGAGCAGGTGGCAGCAGGGGAACGTCTTCTCATGCTC<br>CGTGATGCATGAGGCTCTGCACAACCACTACACGCAGAAGAGCCTCTCCCTGTCTC<br>CGGGTAAA |

| | |
|---|---|
| **IgG3 heavy chain** | caggtccagctgcagcagtctggacctgagctggtgaagcctggggcctcagtgaagatgtcctgcaaggcttctgg<br>ctacaccttcacaagctaccatatacagtgggtgaagcagaggcctggacagggacttgagtggattggatggattta<br>tcctggagatggtagtactcagtacaatgagaagttcaagggcaagaccacactgactgcagacaaatcctccagcac<br>agcctacatgttgctcagcagcctgacctctgaggactctgcgatctatttctgtgcaagggaggggacctactatgct<br>atggactactggggtcaaggaacctcagtcaccgtctcctcaGCTAAAACAACACCCCCATCAGTCTA<br>TCCACTGGCCCCTGGGTGTGGAGATACAACTGGTTCCTCTGTGACTCTGGGATGCC<br>TGGTCAAGGGCTACTTCCCTGAGTCAGTGACTGTGACTTGGAACTCTGGATCCCTGT<br>CCAGCAGTGTGCACACCTTCCCAGCTCTCCTGCAGTCTGGACTCTACACTATGAGC<br>AGCTCAGTGACTGTCCCCTCCAGCACCTGGCCAAGTCAGACCGTCACCTGCAGCGT<br>TGCTCACCCAGCCAGCAGCACCACGGTGGACAAAAAACTTGAATTGAAGACACCT<br>CTCGGCGACACTACTCACACATGCCCAAGATGCCCAGAGCCTAAGTCCTGCGACA<br>CCCCTCCTCCCTGTCCTAGATGCCCTGAGCCAAAGTCTTGCGATACGCCTCCACCCT<br>GCCCTCGGTGTCCTGAGCCTAAATCATGCGATACCCCACCACCATGTCCTCGCTGC<br>CCCGCACCTGAACTCCTGGGAGGACCGTCAGTCTTCCTCTTCCCCCCAAAACCCAA<br>GGATACCCTTATGATTTCCCGGACCCCTGAGGTCACGTGCGTGGTGGTGGACGTGA<br>GCCACGAAGACCCCGAGGTCCAGTTCAAGTGGTACGTGGACGGCGTGGAGGTGC<br>ATAATGCCAAGACAAAGCCGCGGGAGGAGCAGTACAACAGCACGTTCCGTGTGGT<br>CAGCGTCCTCACCGTCCTGCACCAGGACTGGCTGAACGGCAAGGAGTACAAGTGC<br>AAGGTCTCCAACAAAGCCCTCCCAGCCCCCATCGAGAAAACCATCTCCAAAACCAA<br>AGGACAGCCCCGAGAACCACAGGTGTACACCCTGCCCCCATCCCGGGAGGAGAT<br>GACCAAGAACCAGGTCAGCCTGACCTGCCTGGTCAAAGGCTTCTACCCCAGCGAC<br>ATCGCCGTGGAGTGGGAGAGCAGCGGGCAGCCGGAGAACAACTACAACACCACG<br>CCTCCCATGCTGGACTCCGACGGCTCCTTCTTCCTCTACAGCAAGCTCACCGTGGA<br>CAAGAGCAGGTGGCAGCAGGGGAACATCTTCTCATGCTCCGTGATGCATGAGGCT<br>CTGCACAACCGCTTCACGCAGAAGAGCCTCTCCCTGTCTCCGGGTAAA |

| | |
|---|---|
| **IgG4 heavy chain** | caggtccagctgcagcagtctggacctgagctggtgaagcctggggcctcagtgaagatgtcctgcaaggcttctgg<br>ctacaccttcacaagctaccatatacagtgggtgaagcagaggcctggacagggacttgagtggattggatggattta<br>tcctggagatggtagtactcagtacaatgagaagttcaagggcaagaccacactgactgcagacaaatcctccagcac<br>agcctacatgttgctcagcagcctgacctctgaggactctgcgatctatttctgtgcaagggaggggacctactatgct<br>atggactactggggtcaaggaacctcagtcaccgtctcctcaGCTAAAACAACACCCCCATCAGTCTA<br>TCCACTGGCCCCTGGGTGTGGAGATACAACTGGTTCCTCTGTGACTCTGGGATGCC<br>TGGTCAAGGGCTACTTCCCTGAGTCAGTGACTGTGACTTGGAACTCTGGATCCCTGT<br>CCAGCAGTGTGCACACCTTCCCAGCTCTCCTGCAGTCTGGACTCTACACTATGAGC<br>AGCTCAGTGACTGTCCCCTCCAGCACCTGGCCAAGTCAGACCGTCACCTGCAGCGT<br>TGCTCACCCAGCCAGCAGCACCACGGTGGACAAAAAACTTGAGTCCAAATATGGT<br>CCCCCATGCCCATCATGCCCAGCACCTGAGTTCCTGGGGGGACCATCAGTCTTCCT<br>GTTCCCCCCAAAACCCAAGGACACTCTCATGATCTCCCGGACCCCTGAGGTCACGT<br>GCGTGGTGGTGGACGTGAGCCAGGAAGACCCCGAGGTCCAGTTCAACTGGTACGT<br>GGATGGCGTGGAGGTGCATAATGCCAAGACAAAGCCGCGGGAGGAGCAGTTCAA<br>CAGCACGTACCGTGTGGTCAGCGTCCTCACCGTCCTGCACCAGGACTGGCTGAAC<br>GGCAAGGAGTACAAGTGCAAGGTCTCCAACAAAGGCCTCCCGTCCTCCATCGAGA<br>AAACCATCTCCAAAGCCAAAGGGCAGCCCCGAGAGCCACAGGTGTACACCCTGCC<br>CCCATCCCAGGAGGAGATGACCAAGAACCAGGTCAGCCTGACCTGCCTGGTCAAA<br>GGCTTCTACCCCAGCGACATCGCCGTGGAGTGGGAGAGCAATGGGCAGCCGGAG<br>AACAACTACAAGACCACGCCTCCCGTGCTGGACTCCGACGGCTCCTTCTTCCTCTA<br>CAGCAGGCTCACCGTGGACAAGAGCAGGTGGCAGGAGGGGAATGTCTTCTCATGC<br>TCCGTGATGCATGAGGCTCTGCACAACCACTACACACAGAAGAGCCTCTCCCTGTC<br>TCCGGGTAAA |

Lowercase: $V_L$ and $V_H$
Underlined: hinge region
Red: Murine origin
Black: Human part origin

**Supplemental Table 4. Details of HLA molecules used for the purification of HLA-specific IgGs.** Purifications using both beadsets showed comparable MFI values, ensuring that the binding of IgGs was specific to the HLA molecules and not to the loaded peptides. Purifications of all the cohort samples were done using the monomers in bold.

| Specificity | Allele | Loaded peptide | Origin |
|---|---|---|---|
| HLA-A1 | HLA A*01:01 | **VTEHDTLLY** | CMV, pp50 |
| | | CTELKLSDY | Influenza A virus, Nucleoprotein, 44-52 |
| HLA-A2 | HLA A*02:01 | **CLGGLLTMV** | EBV membrane protein, 42-434 |
| | | GLCTLVAML | EBV, mRNA export factor ICP27, 300-308 |
