## Supplemental Figures for "Afucosylation of HLA-specific IgG1 as a potential predictor of antibody pathogenicity in kidney transplantation"

| Supplemental Figures | |
|---|---|
| Supplemental Figure 1 | mAb controls for subclassing assay |
| Supplemental Figure 2 | Relationship between HLA-A2 specific IgG signal and signal reported from the clinical testing |
| Supplemental Figure 3 | Schematic of affinity purification and Fc-glycoprofiling glycosylation analysis of HLA-A2 specific antibodies |
| Supplemental Figure 4 | Proof of concept for antigen specific antibody purification |
| Supplemental Figure 5 | Antigen specific antibody purification profiles |
| Supplemental Figure 6 | Schematic of determination of Antibody-Receptor dissociation kinetics by Biolayer Interferometry (BLI) |
| Supplemental Figure 7 | FcγRI binding characterization of unmodified and afucosylated HLA-A2 IgG1 and IgG3 mAbs |
| Supplemental Figure 8 | FcγRIIIA V158 signaling characterization of polyclonal serum HLA-A2-specific antibodies |
| Supplemental Figure 9 | HLA-A2 specific DSA characteristics in individuals with and without AMR. |

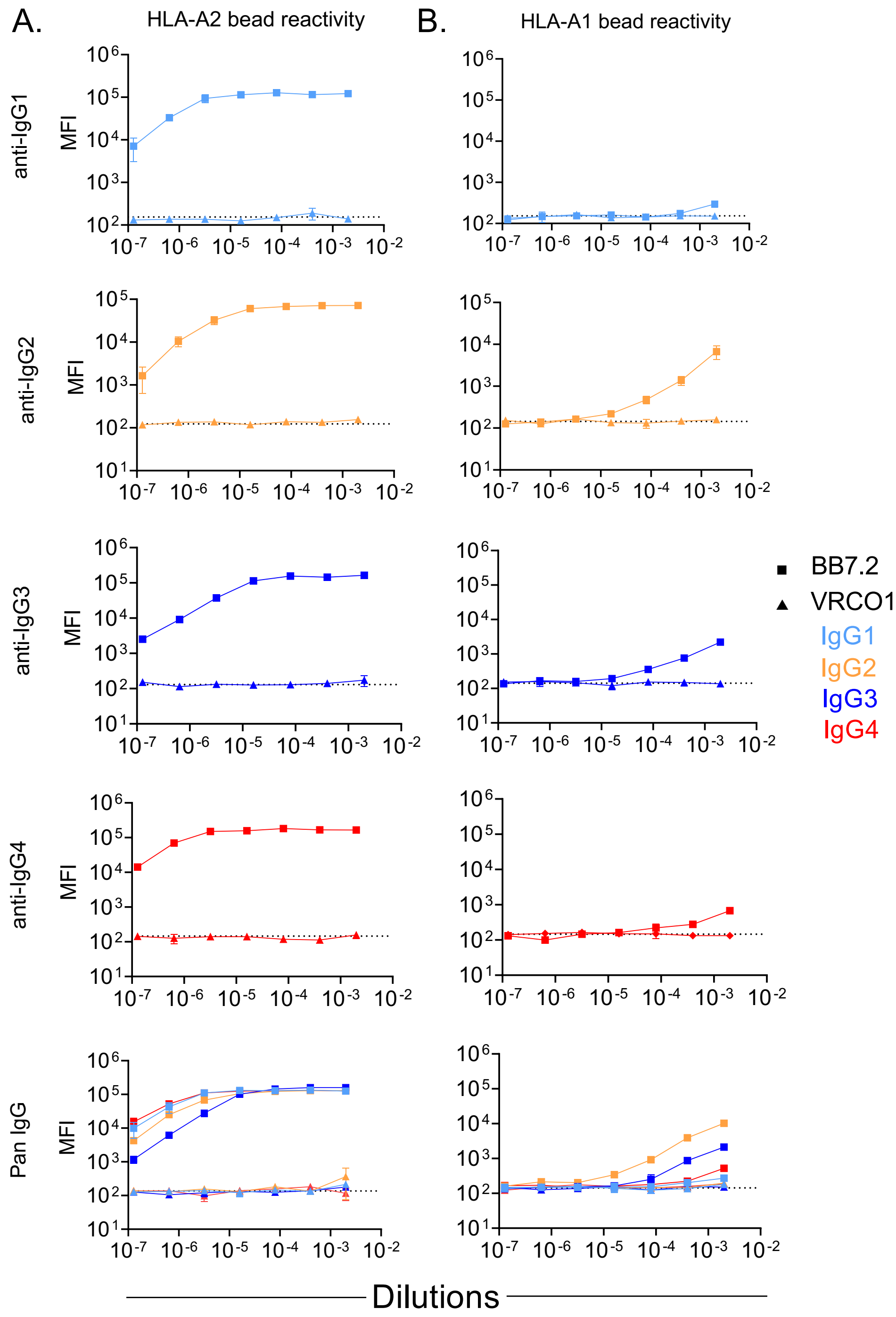


**Supplemental Figure 1. mAb controls for subclassing assay. A-B.** Dose response profiles of HLA-A2 (positive control) and VRCO1 (negative control) mAbs to HLA-A2 (**A**) and HLA-A1 (**B**) antigens across IgG subclasses. HLA-A2-specific BB7.2 (square) and HIV-specific VRCO1 (triangle) mAb subclass controls are shown for each subclass; IgG1 (light blue), IgG2 (orange), IgG3 (dark blue) and IgG4 (red). Baseline (buffer only control) signal is indicated by dotted lines.

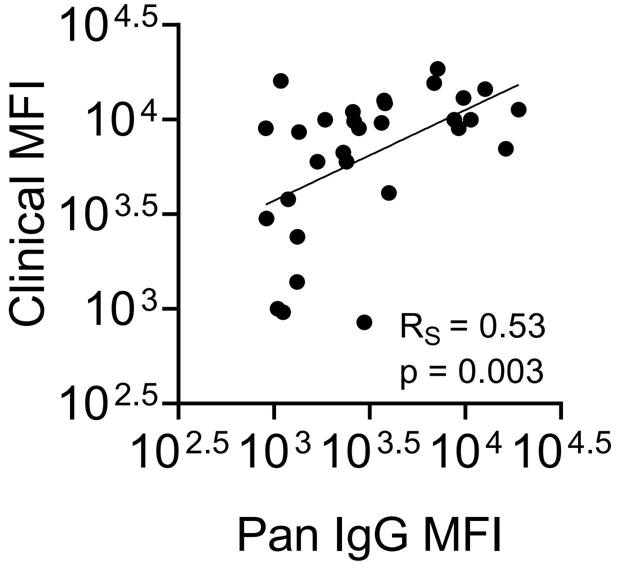


**Supplemental Figure 2. Relationship between HLA-A2 specific IgG signal and signal reported from the clinical testing.** Correlation between IgG titer and values reported from the clinical testing (n=30). Spearman rank correlation ($R_S$) and corresponding *p*-value are shown in inset.

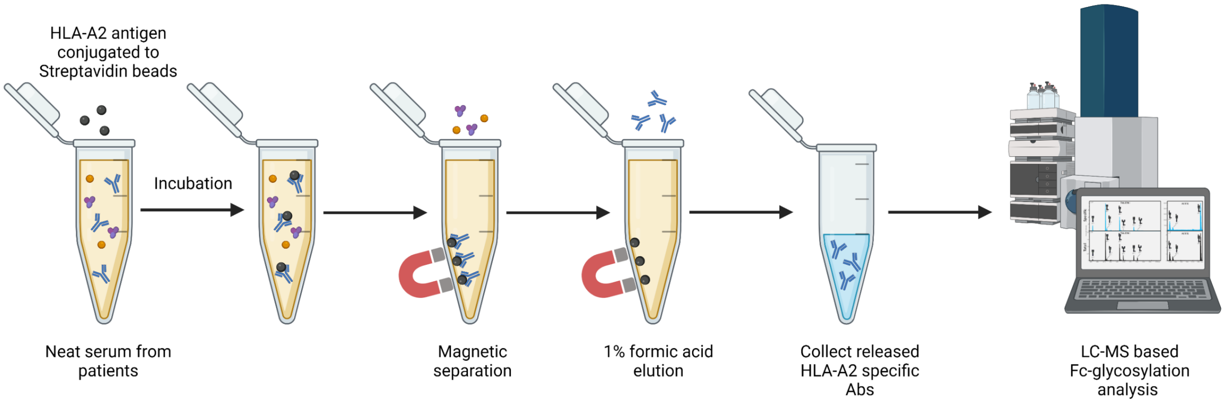


**Supplemental Figure 3. Schematic of affinity purification and Fc-glycosylation analysis of HLA-A2 specific antibodies.** HLA-A2 specific antibodies were purified using HLA-A2 antigen coated magnetic beads. Diluted serum was incubated with magnetic beads to allow binding of HLA-A2-specific antibodies. Beads were washed and then the bound antibodies were eluted. Fc-glycosylation analysis of the eluted HLA-A2 specific antibodies was performed by liquid chromatography – mass spectrometry on the glycopeptide level following tryptic digestion. This figure was created with http://BioRender.com.

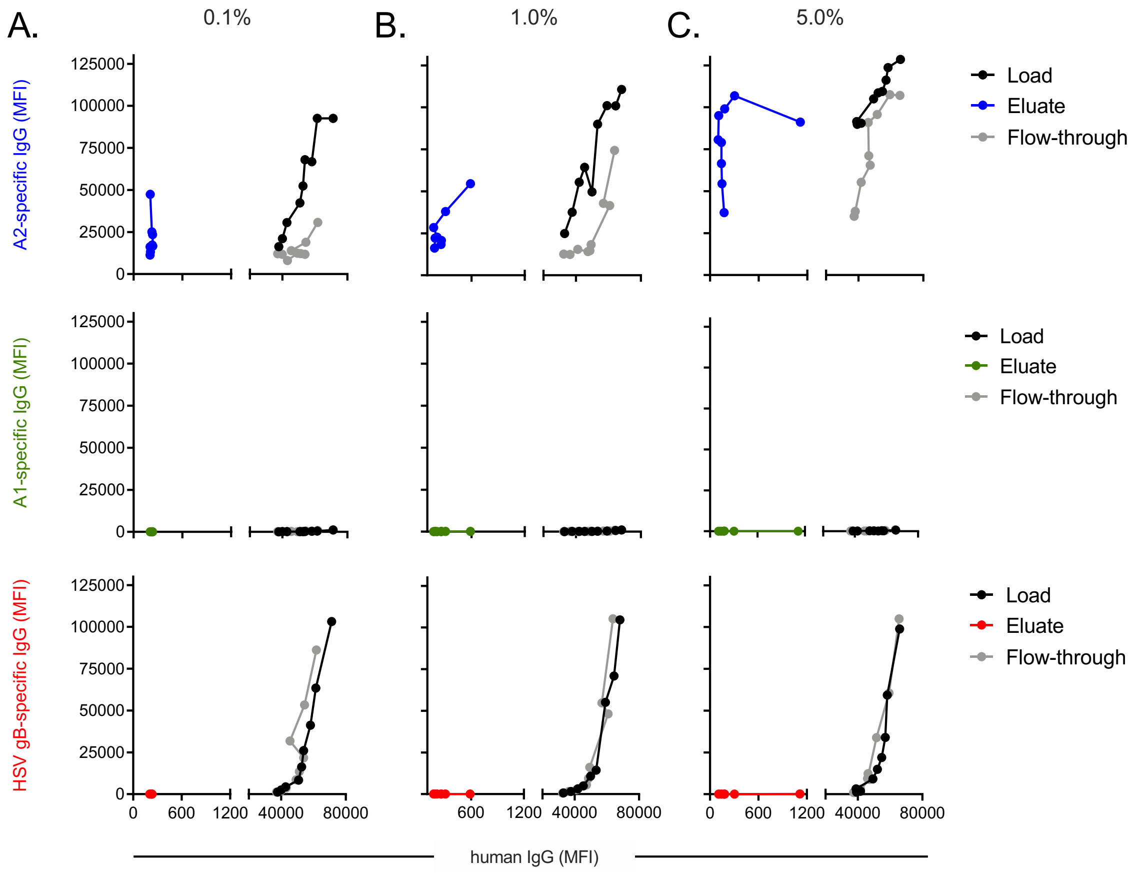


**Supplemental Figure 4. Proof of concept for antigen specific antibody purification. A-C.** Reactivity of IVIG spiked with 0.1% (**A**), 1.0% (**B**) and 5.0% (**C**) murine HLA-A2 mAb to HLA-A2 (top), HLA-A1 (middle) and HSV-gD (bottom) antigens. Load and Flow-through are shown in black and grey, respectively. Reactivity of Eluate for HLA-A2, HLA-A1 and HSV-gD are shown in blue, green, and red, respectively.

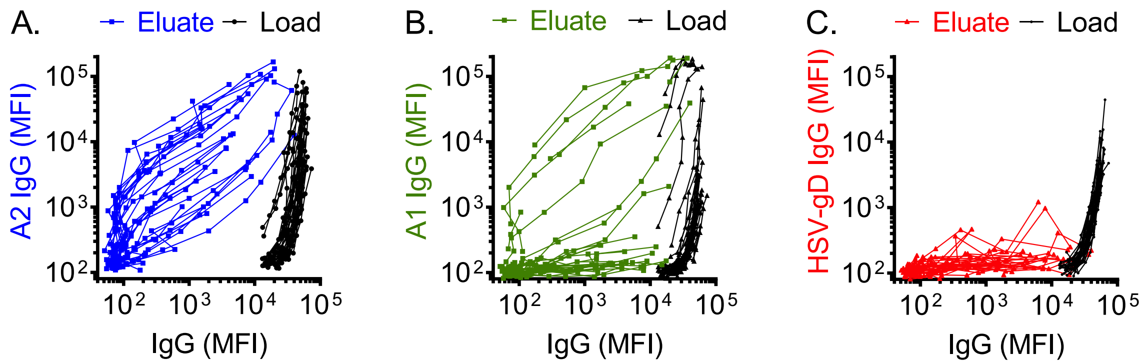


**Supplemental Figure 5. Antigen specific antibody purification profiles. A-C.** Reactivity of purified HLA-A2 specific antibodies to HLA-A2 (**A**), HLA-A1 (**B**) and HSV-gD (**C**) antigens from HLA-A2 positive (n=29) individuals. Signal for each antigen specificity in plotted on the y-axis and signal from total IgG on the x-axis. Reactivity of antibodies in eluate to HLA-A2, HLA-A1 and HSV-gD antigens are shown in blue, green and red, respectively. Profiles of the serum antibodies before purification (loads) are shown in black.

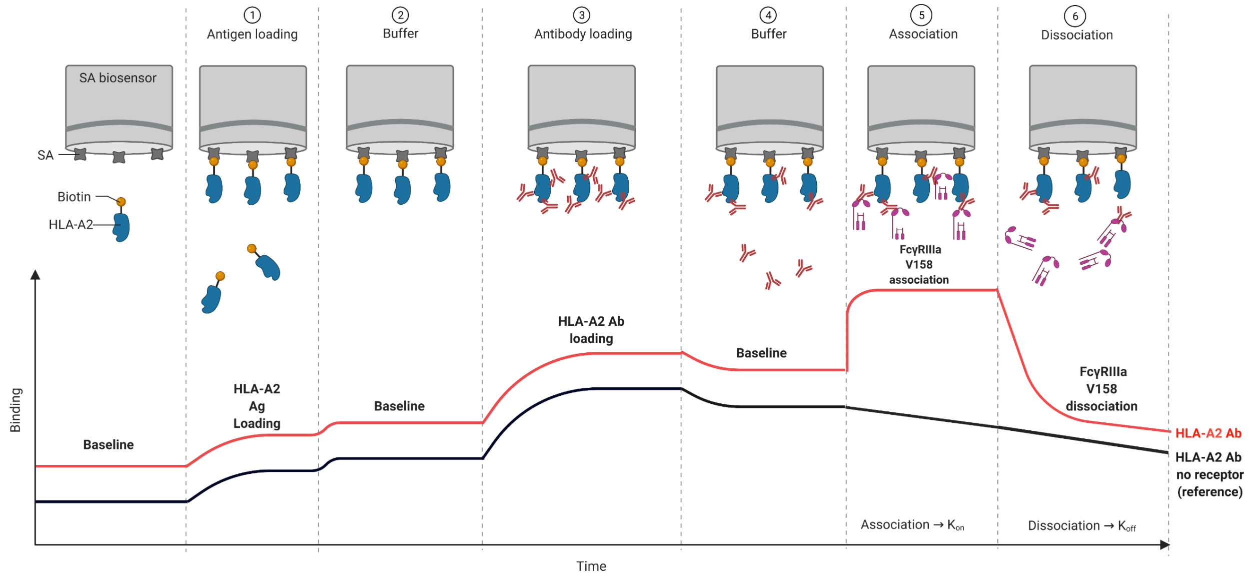


**Supplemental Figure 6. Schematic of the Antibody-Receptor dissociation kinetics measurement by Biolayer Interferometry (BLI).** Streptavidin tips were first coated with biotinylated HLA-A2 antigens. After establishing a signal baseline, the tips were loaded with HLA-A2-specific antibodies, dipped into the buffer for baseline, and then finally dipped into a solution of recombinantly expressed FcγRIIIa V158 monomers. Following receptor association, tips were dipped into a buffer, which allowed receptor dissociation. The dissociation rate of FcγR was defined relatively to a reference tip which was not dipped into the receptor. This figure was created with http://BioRender.com.

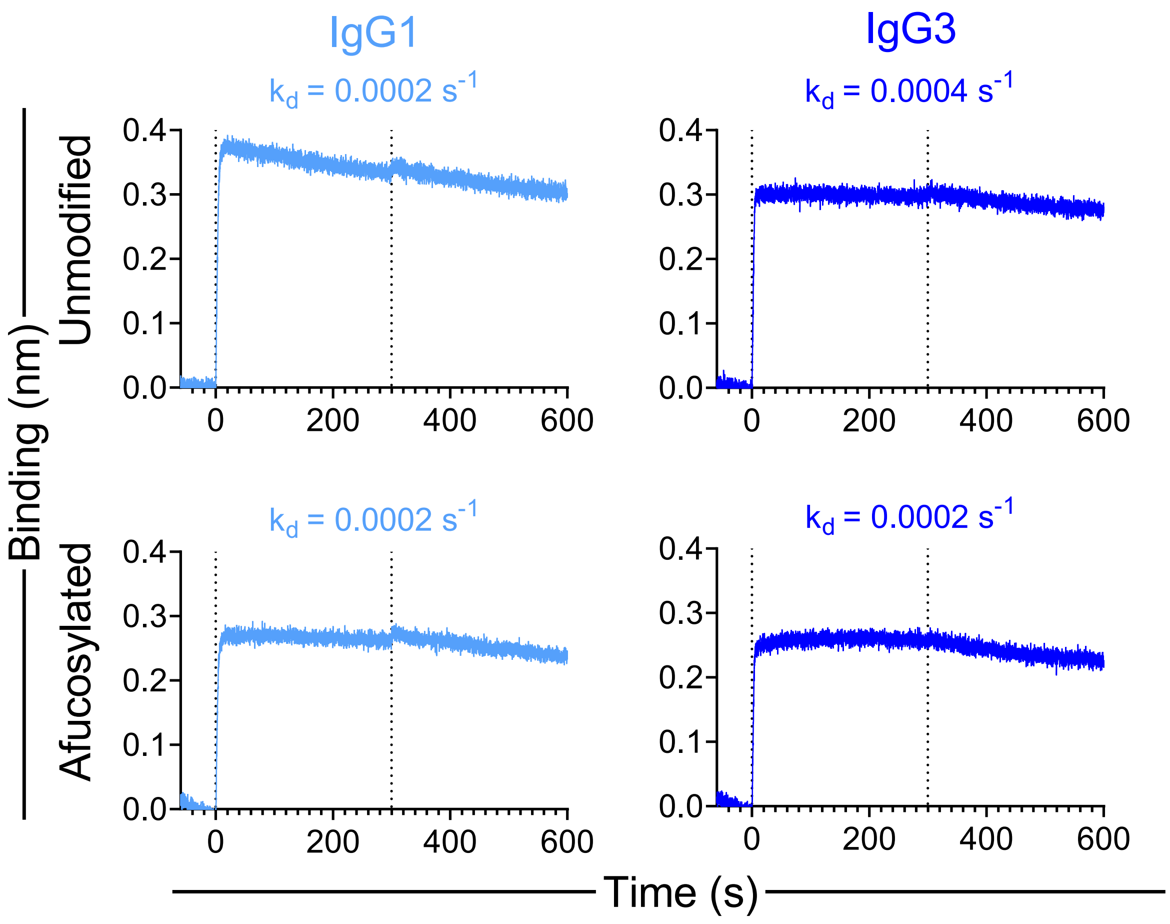


**Supplemental Figure 7. FcγRI binding characterization of unmodified and afucosylated HLA-A2 IgG1 and IgG3 mAbs.** FcγRI association with and dissociation from unmodified and afucosylated HLA-A2 mAbs. Dissociation rates ($k_d$) are shown in inset.

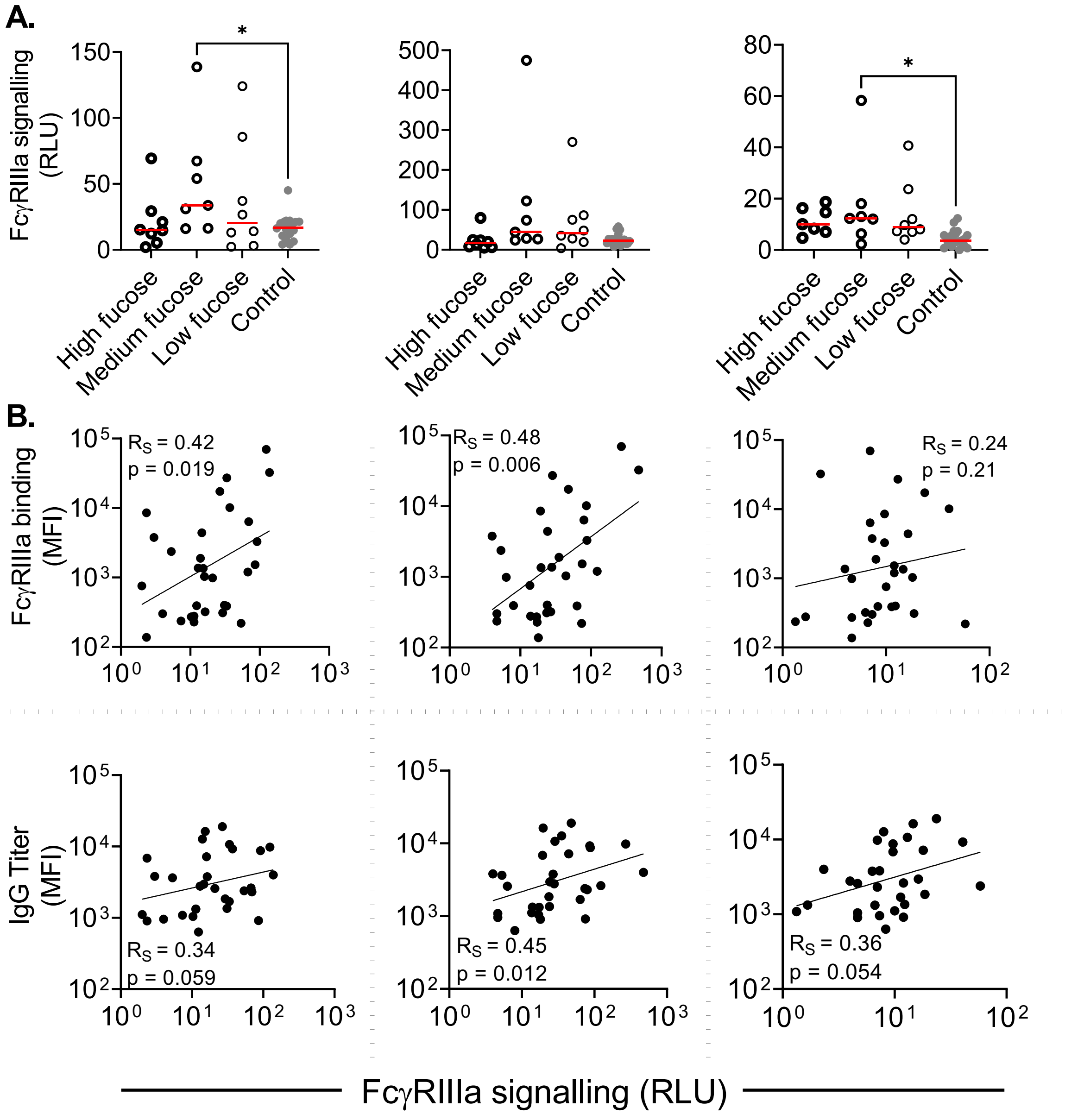


**Supplemental Figure 8. FcγRIIIa V158 signaling characterization of polyclonal serum HLA-A2 specific antibodies. A.** Fc**γ**RIIIA signaling in a reporter cell line assay with high (n=7-8), medium (n=7) and low (n=8) IgG1 fucose content, and controls (n=18) in three independent assay replicates. ADCC assay performed with direct antigen (left and middle), and neutravidin-antigen (right) coated on the plate. Statistical analysis was performed using Ordinary one-way ANOVA adjusted for multiple comparisons using Tukey's test (**p*<0.05). **B.** Correlations between Fc**γ**RIIIa signaling to Fc**γ**RIIIa binding (middle panel) and IgG titer (lower panel) (n=30-31) across Fc signaling assay replicates. Spearman rank correlations ($R_S$) are shown in inset.

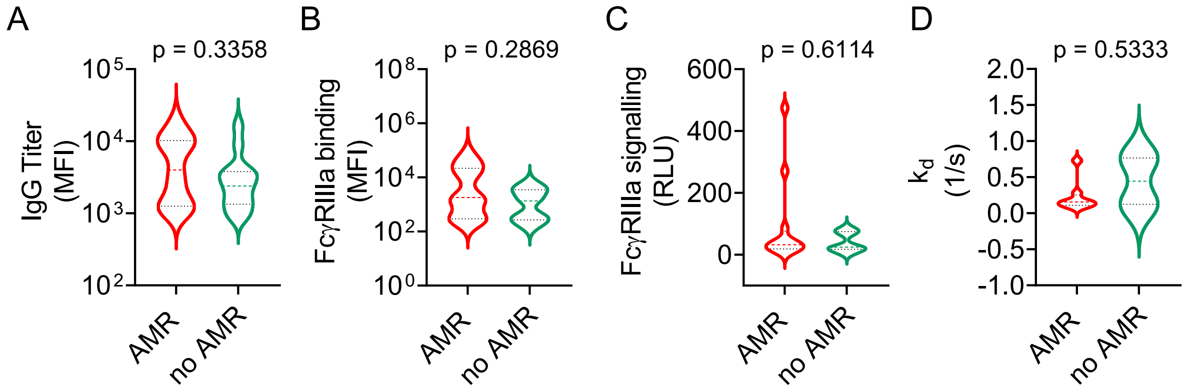


**Supplemental Figure 9. HLA-A2-specific antibody characteristics in individuals with and without AMR. A-D.** Violin plots showing HLA-A2-specific IgG titer in individuals with AMR (n=13) versus no AMR (n=13) **(A)**, Fc**γ**RIIIa binding characterization in individuals with AMR (n=13) versus no AMR (n=13) **(B)**, Fc**γ**RIIIa signaling characterization in individuals with AMR (n=12) versus no AMR (n=13) **(C)** and Fc**γ**RIIIa dissociation rate in individuals with AMR (n=8) versus no AMR (n=2) **(C)**. Patients with and without AMR are shown in red and green, respectively. Statistical analysis was performed using a Mann-Whitney U test.
